# Peripheral inflammation is associated with structural brain atrophy and cognitive decline linked to mild cognitive impairment and Alzheimer’s disease

**DOI:** 10.1101/2023.12.08.23299734

**Authors:** Nuanyi Liang, Kwangsik Nho, John W. Newman, Matthias Arnold, Kevin Huynh, Peter J. Meikle, Kamil Borkowski, Rima Kaddurah-Daouk, Alzheimer’s Disease Metabolomics Consortium, Alzheimer’s Disease Neuroimaging Initiative

## Abstract

Mounting evidence points towards inflammation as an important factor in Alzheimer’s disease (AD) pathogenesis. In this study, we investigated the relationship between a key marker of inflammation – glycoprotein acetyls (GlycA) – in blood and cognitive and brain structural changes in over 1500 participants enrolled into the Alzheimer’s Disease Neuroimaging Initiative. We evaluated those associations cross-sectionally at baseline, followed by an evaluation of whether baseline GlycA can inform about future disease progression. Our results support the following findings: 1) GlycA is elevated in participants diagnosed with AD compared to cognitively normal participants; 2) GlycA level correlates negatively with regional brain volumes in females diagnosed with late mild cognitive impairment (LMCI) or AD; 3) baseline GlycA level is associated with executive function decline at 3-9 year follow-up in both male and female participants diagnosed with LMCI at baseline; and 4) baseline GlycA is associated with decline in entorhinal cortex volume at years 2, 4 and 6-8 of follow-up in both male and female participants diagnosed with LMCI at baseline. In conclusion, peripheral inflammation was found to be positively associated with AD diagnosis and future decline in cognition and regional brain volumes. However, we note that the cross-sectional relationship between peripheral inflammation and AD-related brain atrophy is specific to sex and diagnostic status. Our findings point to peripheral inflammation as a risk factor in AD development, which enables the identification of potential markers and therapeutic intervention for participants who are at risk.

## INTRODUCTION

Mounting evidence suggests that inflammation and immune dysregulation play a critical role in Alzheimer’s disease (AD) pathogenesis (1). Inflammatory-related disorders such as metabolic syndromes, diabetes, and cardiovascular diseases are linked to an increased risk of AD (2–6). In addition, disturbances of circulating markers of inflammation, such as cytokines, endocannabinoids, and oxylipins, have been linked to AD and cognitive decline (7–10). In pre-clinical models, the peripheral inflammation stimuli, such as a high-fat diet and chronic lipopolysaccharide administration, can lead to AD-related dysregulations in the central nervous system (CNS), including altered blood-brain barrier functionality, changed cerebrovascular properties, neuroinflammation, and amyloid beta (Aβ) pathologies (11–16). In addition, factors such as sex differences (17, 18) and health status (19, 20) may alter the crosstalk between peripheral inflammation and brain pathologies. However, despite the linkage of peripheral metabolism to central pathologies (13, 21–23), we still need a better understanding of the crosstalk between peripheral inflammation and AD characteristics in the CNS to guide the improvement of inflammatory-related modifiable actions for reducing AD risks and/or untoward pathological trajectories.

Glycoprotein acetyls (GlycA), a biomarker for systemic inflammation, is informative for vascular aging (24), cardiovascular risk (25–27) and mortality (28). This proton nuclear magnetic resonance (NMR)-based measurement detects the production and further glycosylation of several acute-phase proteins, including alpha-1-acid glycoprotein, alpha-1-antitrypsin, alpha-1-antichymotrypsin, haptoglobin and transferrin (29). As such, GlycA levels capture the overall burden of both the acute and chronic phases of inflammation (26, 29), which has been indicative for inflammatory disorders such as rheumatoid arthritis (30, 31), lupus (32, 33), psoriasis (34), atherosclerotic cardiovascular diseases (26, 27), metabolic syndrome (25, 35), type 2 diabetes (36, 37), and inflammatory bowel disease (38). Compared to the established inflammatory biomarker C-reactive protein (CRP), GlycA may have the advantage of reduced biological variability, as a previous study has shown that the weekly intraindividual and interindividual variations of 23 healthy participants over 5 weeks were substantially lower for GlycA (4.3% and 15.3%, respectively) than for CRP (29.2% and 133.9%, respectively) (29). Relations between plasma GlycA and cognitive functions has been reported in a study of young healthy adults (39), but such a study is not entirely informative for neurodegenerative diseases that are usually associated with old ages. Therefore, the relationship between GlycA and AD-related biomarkers in older participants has yet to be established.

To further address these knowledge gaps, we aimed in this study to determine the relationship between peripheral inflammation and AD-related biomarkers. We evaluated: 1) differences in GlycA levels in participants at different diagnosis stages along the AD continuum; 2) baseline GlycA correlation with longitudinal brain regional brain volumes, memory and executive function at baseline and in the follow-up years; and 3) the influence of diagnosis status and sex on those correlations. To do so, we utilized the data from the Alzheimer’s Disease Neuroimaging Initiative (ADNI) cohort, including baseline NMR measurement of GlycA, baseline and longitudinal magnetic resonance imaging (MRI) of brain morphologies, executive function composite score, and memory composite scores to perform correlation analysis.

## METHODS

### ADNI Study Participants

Baseline demographic, clinical, cognition, MRI imaging, genetic, biomarkers, and multi-omics data of the study participants used in the current study were acquired from the ADNI database (www.adni-info.org), managed through the Laboratory of Neuro Imaging Image & Data Archive (http://adni.loni.usc.edu/). The protocol procedures were approved by the Institutional Review Boards or Research Ethics Boards of the participating institutions. Participants provided signed written informed consent and permission for data access and analysis. The ADNI cohort, launched in 2003 as a public-private partnership and led by Principal Investigator Michael W. Weiner, MD, tests the primary goal of combining serial MRI and positron emission tomography (PET) imaging, biological markers, and clinical and neuropsychological evaluation to measure the progression of mild cognitive impairment (MCI) and early AD.

The diagnosis status of the adult participants included cognitively normal (CN), significant memory concern (SMC), early mild cognitive impairment (EMCI), late mild cognitive impairment (LMCI), and AD. The detailed inclusion and exclusion criteria can be accessed at the ADNI documentation website (https://adni.loni.usc.edu/methods/documents/). In brief, CN participants had Mini-Mental State Exam (MMSE) scores of 24-30 (inclusive) and a Clinical Dementia Rating scale (CDR) score of 0 without any memory complaints. SMC participants had self-reported subjective memory concerns but MMSE and CDR scores rated as CN (https://classic.clinicaltrials.gov/ct2/show/NCT01231971). MCI participants had an MMSE score of 24-30 (inclusive), a CDR score of 0.5, and a memory complaint, yet did not have sufficient levels of cognition and functional performance impairment to be diagnosed with dementia. Among MCI participants, EMCI and LMCI were differentiated by different levels of abnormal memory functions, which was indicated by the education-adjusted Wechsler Memory Scale-Revised Logical Memory II subscale, with the cut-off value for LMCI being a) ≤8 for 16 or more years of education, b) ≤4 for 8-15 years of education and c) ≤2 for 0-7 years of education. AD participants had an MMSE score of 20-26 (inclusive), a CRD of 0.5 or 1.0, a memory complaint, and their cognition and functional performances met the National Institute of Neurological and Communicative Disorders and Stroke/the Alzheimer’s Disease and Related Disorders Association criteria for probable AD. Exclusion criteria included – but were not limited to – major depression and other significant neurological diseases. Other detailed exclusion criteria can be found in the above-mentioned ADNI study documents website. The dataset contained complete info of GlycA, APOE genotypes, sex, body mass index (BMI), age and education (n=1506). Of the participants, 684 (45.4%) were female; 700 (46.5%) were APOE4 carriers, and 153 (10.2%) were APOE4/4 carriers. In addition, 361 (24.0%) were CN participants, 96 (6.4%) had SMC, 280 (18.6%) had EMCI, 481 (31.9%) had LMCI, and 288 (19.1%) had AD. The age at screening (average ± standard deviation) was 73.1 ± 7.1, baseline BMI was 26.9 ± 4.9, and years of education was 15.9 ± 2.9. The detailed protocols for blood and CSF sample collection and storage can be found on the abovementioned ADNI study documents website.

### Peripheral GlycA Measurement

Peripheral GlycA was measured via the Nightingale Health platform using established protocols (40–42). In brief, serum samples were stored at −80 °C and thawed overnight at +4 °C prior to analysis. The samples were then gently mixed, centrifuged, and mixed with an equal amount of an NMR measurement buffer of 75 mmol/L disodium phosphate in 8:2 H_2_O/D_2_O (pH 7.4) with 0.08% sodium 3-(trimethylsilyl)propionate-2,2,3,3-d_4_ and 0.04% sodium azide (40). The prepared samples were then measured using a Bruker AVANCE III HD 500 MHz spectrometer coupled with a SampleJet cooled robotic sample changer and CryoProbe Prodigy TCI, a cryogenically cooled triple resonance probe head (41). Quantification results were then obtained from the NMR spectral data via an advanced proprietary software of the Nightingale Health platform (42).

In addition, the quantitative result of GlycA was log2-transformed and adjusted with medication data prior to further analyses using an AIC backward stepwise regression method (22, 43). Other confounders, such as age, BMI, sex, education or APOE4 were adjusted as specified in the results.

### Cytokine Analysis of Plasma and Cerebral Spinal Fluid

Cytokines in plasma and cerebral spinal fluid (CSF) were measured using fluorescence-labeled microspheres via a multiplex proteomics method based on the Luminex xMAP platform, with an established threshold of detection and a dynamic range for each analyte (44, 45). The detailed sample preparation was provided by the previous publication, which included steps of sample thawing, introduction of capture microspheres, the addition of multiplexed mixtures of reporter antibodies (biotinylated), multiplex development via the streptavidin-phycoerythrin solution, and final volume adjustment (45). Prior to statistical analysis, cytokines were normalized and medication-adjusted as above.

### MRI Brain Imaging Data Acquisition

MRI protocol and data were accessed through the ADNI website (https://adni.loni.usc.edu/). In brief, MRI imaging was acquired using a 1.5T or 3T MRI scanner with the sequences of T1 and dual echo T2-weighted imaging (ADNI 1), or a 3T MRI scanner with fully sampled and accelerated T1-weighted imaging in addition to 2D FLAIR and T2*-weighted imaging (ADNI GO/2).

### Cognitive Function and Memory Measurement

A composite score for measuring overall executive functioning (EF) was previously developed and validated (46). In brief, the composite score was constructed via an iterative process that included the confirmatory factor analysis to build a model and the reviewing of findings to construct a revised model. The model was built using ADNI baseline data, based on the Category fluency (animals) test, Category fluency (vegetables) test, Trails A and Trails B test, Digit span backwards test, Wechsler Adult Intelligence Scale – Revised Digit Symbol Substitution test, and the five Clock Drawing items test (circle, symbol, numbers, hands, and time).

A composite score for measuring the overall memory (MEM) was previously developed and validated (47). In brief, MEM was derived from two versions of the longitudinal Rey Auditory Verbal Learning Test, three versions of the AD Assessment Schedule - Cognition, the MMSE, and Logical Memory data, using a single factor model.

### Amyloid/Tau/Neurodegeneration (A/T/N) Biomarker Measurement

Amyloid/Tau/Neurodegeneration (A/T/N) biomarker, Aβ_1-42_, t-tau, and p-tau_181p_ were measured via a fully automated Roche Elecsys immunoassay, in which a sample preparation procedure with two incubation steps was used (48, 49).

### Statistical Analyses

All statistical analyses were performed in JMP® Pro 16 and 17 (SAS Institute Inc., Cary, NC). We used the standard least-square method for analysis of covariance (ANCOVA) or calculating the residual of variable controlling confounders. ANCOVA was applied to compare the differences in means. To investigate non-linear associations, Spearman’s correlations were performed on residuals of specific confounders, namely APOE4, age, BMI, education level, sex, diagnosis status and/or medications. For the MRI measurement of brain volumes, we adjusted for the confounders of magnet type of MRI and intracranial volume (log2-transformed). A linear model with sex or diagnosis status interaction with GlycA was additionally applied to test whether the observed associations were different between sexes or among diagnosis status.

Additional Spearman’s correlation analyses were performed to evaluate the relationship of baseline GlycA with cognitive functions and brain regional volumes in the future follow-up years with the same confounders adjusted as above, in addition to adjusting to the baseline value of these measurements. The analysis was performed with stratification of diagnosis status at baseline, because participants with differing diagnosis status at baseline experienced different trajectories of brain structural and cognitive changes in the follow-up years (**Figure 1**).

**Figure 1.**
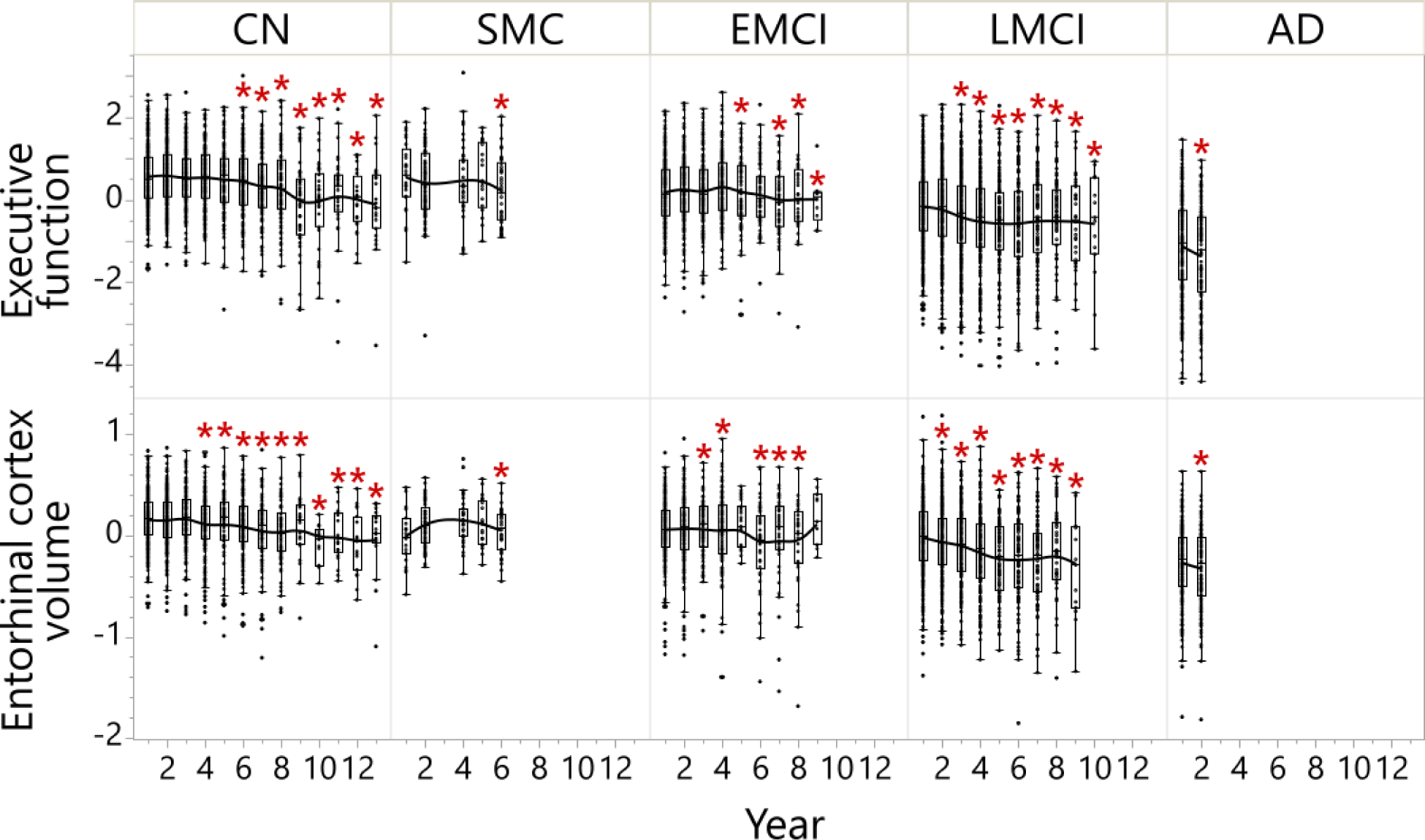
Participants diagnosed with LMCI at baseline started to have the most continuous executive functional decline on and after 3-year follow-up and continuous entorhinal cortex volume decline on and after 2-year follow-up, which was well distinguished from CN, SMC, EMCI and AD participants. **Executive function** was executive function score adjusted for sex, ApoE4, education, BMI at visit and age at screening. The ANCOVA results comparing different time points within each diagnosis group are shown in the heatmap of **Figure S3**; the ANCOVA results comparing EF of different diagnosis at baseline are shown in **Figure S4**. **Entorhinal cortex volume** was entorhinal cortex volume adjusted for intracranial volume (log2), magnet type, sex, ApoE4, education, BMI at visit and age at screening. The ANCOVA results comparing different time points within each diagnosis group are shown in the heatmap of **Figure S7**; the ANCOVA results comparing the entorhinal cortex volume of different diagnosis at baseline are shown in Figure 5. Results annotated with * were at a significantly different level compared to Year 1 within the diagnosis group (**Figure S3** and **S7**). *Abbreviations:* AD: Alzheimer’s disease; ANCOVA: Analysis of covariance; BMI: Body mass index; CN: Cognitively normal; EMCI: Early mild cognitive impairment; LMCI: Late mild cognitive impairment; SMC: Significant memory concerns.

## RESULTS

### Inflammation Level Indicated by GlycA was Elevated along the AD Trajectory

To evaluate the level of peripheral inflammation along AD progression, we applied an ANCOVA model that tested GlycA level against diagnosis groups (CN, SMC, EMCI, LMCI and AD) and controlled for the presence of APOE4 allele, age, BMI, education and sex as covariates (**Figure 2a**). The sex-diagnosis group interaction was additionally applied to test whether sex influences the pattern of GlycA levels along the disease progression and it was not significant (*p* = 0.579).

**Figure 2.**
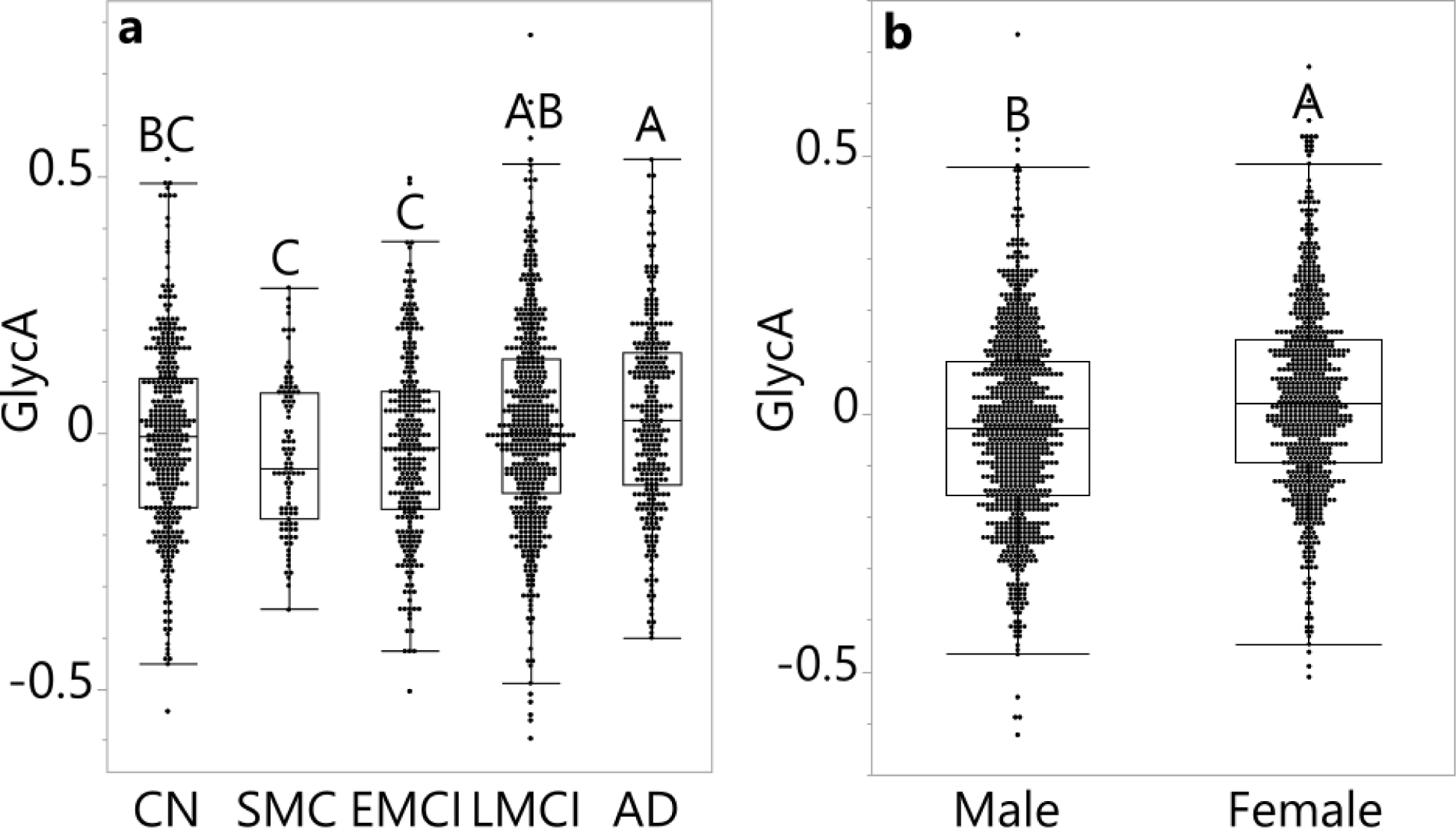
GlycA level in a) groups of various diagnosis status, adjusted for sex and b) sexes, adjusted by diagnosis status. **a)** GlycA level was log2-transformed and adjusted for medications, sex, age, APOE4, BMI, and education level. **b)** GlycA level was log2-transformed and adjusted for medications, diagnosis status, age, APOE4, BMI, and education level. ANCOVA and Tukey HSD post hoc analysis were performed; levels not labelled with the same letter differed significantly (*p* <0.05). Values in the bar graph were shown as the mean residual GlycA level ± standard error. There was no sex-diagnosis status interaction in a full factorial model. *Abbreviations:* AD: Alzheimer’s disease; ANCOVA: Analysis of covariance; BMI: Body mass index; CN: Cognitively normal; EMCI: Early mild cognitive impairment; GlycA: Glycoprotein acetyls; HSD: Honestly significant difference; LMCI: Late mild cognitive impairment; SMC: Significant memory concerns.

GlycA level increased in the participants at more severe disease stages (diagnosis group *p*<0.001). In particular, AD participants had higher GlycA levels compared to participants with CN, SMC and EMCI. Participants with LMCI had higher GlycA levels than did participants with SMC and EMCI. In the same model, GlycA was positively associated with BMI (*p*<0.001, β_estimate_=0.01) and negatively associated with education (*p*<0.001, β_estimate_=-0.01). GlycA was also higher in females than in males (*p*<0.001, d_Cohen_=0.3, **Figure 1b**). GlycA was not influenced by the presence of APOE4 (*p*=0.227) or age (*p*=0.143).

Additionally, we evaluated the relationship between plasma GlycA and plasma CRP using Spearman’s rank order correlation to compare the NMR-derived inflammation indicator to the commonly used peripheral inflammation marker (**Figure S1A**). Moreover, we correlated plasma GlycA and CRP in CSF to evaluate the association between peripheral and central inflammation (**Figure S1B**). GlycA was positively associated with CRP in both plasma (*p*<0.001, ρ=0.45) and CSF (*p*<0.001, ρ=0.40). In contrast, CRP in both plasma and CSF did not differ among the different diagnosis groups (p=0.192 and p=0.832), and only CRP in plasma was higher in females compared to males (*p*=0.015, d_Cohen_=0.2) (**Figure S2**), where fewer participants had their CRP measured (n for each group: in blood: n=50 (CN), 0 (SMC), 0 (EMCI), 337 (LMCI), and 97 (AD); in CSF: n=74 (CN), 0 (SMC), 0 (EMCI), 133 (LMCI), and 60 (AD)). The lack of significant differences in plasma CRP among diagnosis groups due to reduced data coverage was supported by the result that when adjusting plasma CRP, GlycA levels were no longer significantly different among the various diagnosis groups in the abovementioned ANCOVA model (*p*=0.126).

### Baseline GlycA Level was Correlated with a Future Decrease in Executive Function in Participants Diagnosed with LMCI at Baseline

To interrogate whether baseline GlycA can predict future cognitive decline, we evaluated the correlation of baseline GlycA with the longitudinal composite scores for EF (46). The relationship between baseline GlycA and EF in follow-up years was evaluated using Spearman’s rank order correlation analyses, adjusting for baseline EF, sex, age, APOE4 genotype, BMI, education and follow-up years, and treating participant ID as a random factor. In addition, due to the differential connection observed between central and peripheral changes in different stages of AD progression (50), the analyses were performed with stratification of diagnosis status. A linear mixed model was used to confirm these analyses.

ADNI data with the baseline GlycA measurement enabled us to probe cognitive changes for up to a 13-year follow-up (**Figures 1 and S3**). Participants diagnosed with LMCI at baseline experienced the earliest EF declines (defined as the longitudinal EF significantly deviated from Year 1 follow-up) in follow-up years among participants who were not diagnosed with AD at baseline, starting on Year 3 (**Figures 1 and S3)**. Therefore, we specifically investigated the correlations between baseline GlycA level and EF in the different months of follow-up among participants diagnosed with LMCI at baseline. The LMCI-GlycA interaction to predict EF was confirmed using a linear mixed model (*p*=0.025, **Table S1**). In participants with LMCI, baseline GlycA was negatively associated with EF in years 3-9 of follow-up (**Figure 3**, with year 3 (*p*=0.036, ρ=-0.119), year 4 (*p*=0.001, ρ=-0.235), year 5 (*p*=0.003, ρ=-0.267), year 6 (*p*<0.001, ρ=-0.380), year 7 (*p*=0.002, ρ=-0.314), year 8 (*p*=0.002, ρ=-0.389) and year 9 (*p*=0.040, ρ=-0.345)). Additionally, GlycA-EF association among participants with LMCI was not sex-dependent (*p*_interaction_=0.833) (**Table S2**). The negative correlation between longitudinal EF and baseline GlycA in participants diagnosed with LMCI at baseline was confirmed using a linear mixed model (**Table S3**)

**Figure 3.**
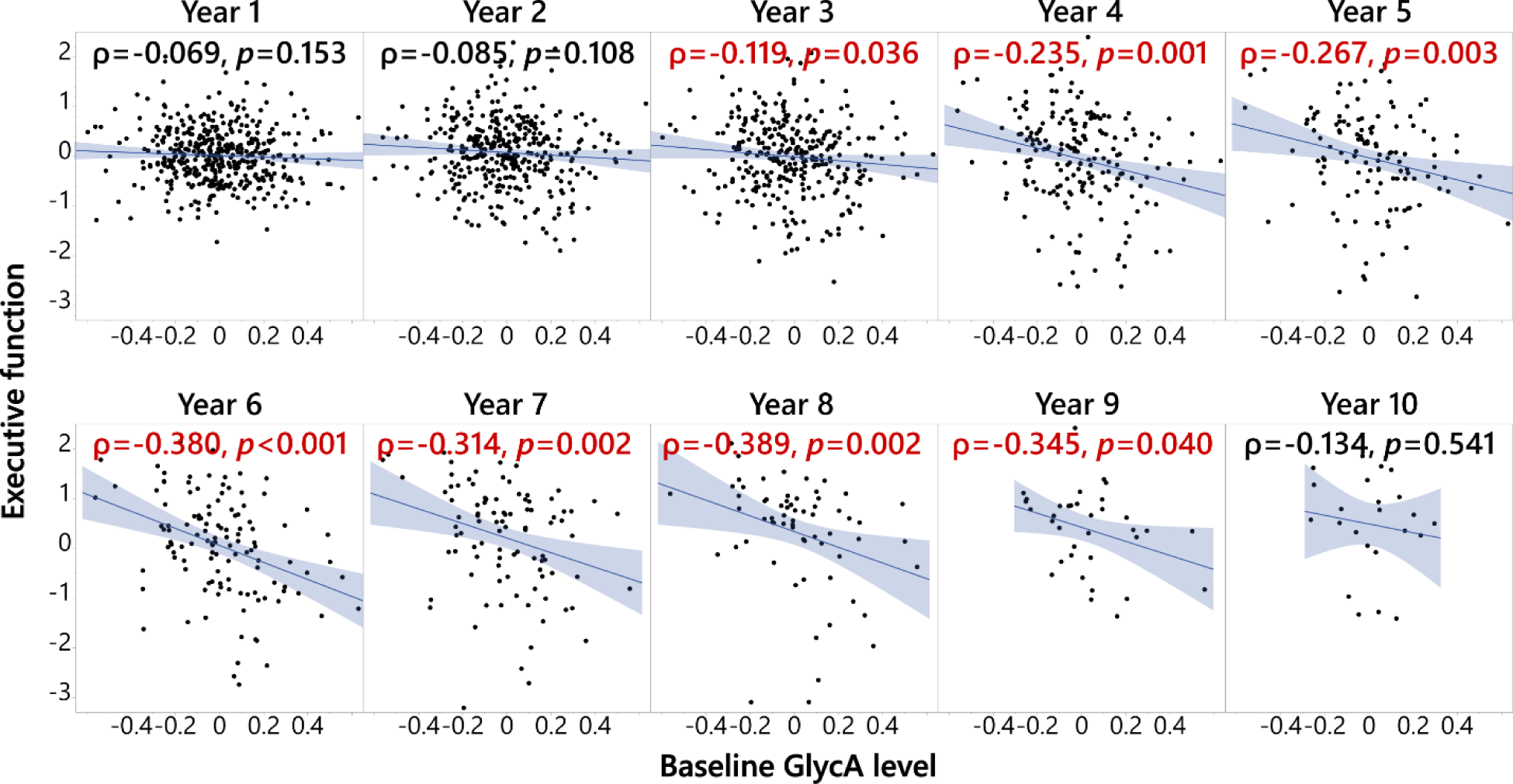
Baseline GlycA level was associated with executive function composite score (EF) in late mild cognitive impairment (LMCI) participants after 3 years of follow-up. Spearman’s rank order correlation was performed on residuals of EF and GlycA, and the analyses were stratified by diagnosis status. EF at different years were adjusted for baseline EF level, screening age, follow-up year, BMI at the time, APOE4, sex, and education, treating participant ID as random factors; baseline GlycA level was adjusted for medication, screening age, baseline BMI, APOE, sex, and education. The analysis was performed on participants with different diagnosis status, but only in the participants with LMCI was EF at continuous follow-up years negatively associated with baseline GlycA level. In addition, the LMCI-GlycA interaction was confirmed using a linear mixed model (**Table S1**). Therefore, the analysis results for the participants with LMCI are shown here. A full factorial linear mixed model was used to show that there were no sex-GlycA interactions controlling DX (*p*=0.864). *Abbreviations:* BMI: Body mass index; DX: Diagnosis at baseline; GlycA: Glycoprotein acetyls; ID: Identification.

Of note, only a weak association (p=0.049, ρ=-0.172) was observed between the baseline GlycA and baseline EF in female participants with AD (**Figure S4**). No associations were detected in females in other diagnostic groups, or in males. Therefore, the lack of sex effects on the GlycA-EF relationship was consistent between cross-sectional and longitudinal observations.

### Baseline GlycA Level was Correlated with a Future Decrease in Memory in Participants Diagnosed with LMCI at Baseline

To investigate whether memory function follows a pattern similar to executive function, similar analyses to the above were performed on baseline GlycA and longitudinal composite scores for MEM (47). Participants diagnosed with LMCI at baseline experienced the earliest continuous cognitive declines (i.e., cognition significantly deviated from Year 1 follow-up) in follow-up years among participants who were not diagnosed with AD at baseline, starting in Year 2 (**Figure S5**). In these participants, baseline GlycA was negatively associated with MEM in years 5-8 of follow-up (**Figure S6**, with year 5 (*p*=0.036, ρ=-0.183), year 6 (*p*=0.037, ρ=-0.186), year 7 (*p*=0.019, ρ=-0.235) and year 8 (*p*=0.036, ρ=-0.262)). However, the associations were weaker compared to associations with EF, which was confirmed using a linear mixed model (**Table S3**). There was also no significant LMCI-GlycA interaction to predict MEM, which was confirmed using a linear mix model (*p*=0.441, **Table S1**). Additionally, GlycA-MEM association among participants with LMCI was not sex-dependent (*p*_interaction_ = 0.471). Unlike baseline EF, there was no association between baseline GlycA and baseline MEM in either sex or in any diagnosis group (*p*>0.05).

### Baseline GlycA Level was Correlated with a Future Decrease in Entorhinal Cortex Volume in Participants Diagnosed with LMCI at Baseline

To identify the structural basis of the MEM and EF patterns observed above, we performed a similar analysis on MRI-measured brain regional volumes with additional adjustment for MRI methods and intracranial volume. We therefore investigated whether baseline GlycA can predict further decline in brain regional volumes using the above-described approach. Participants diagnosed with LMCI at baseline experienced the earliest continuous memory declines (i.e., memory significantly deviated from Year 1 follow-up) in follow-up years among participants who were not diagnosed with AD at baseline, starting in Year 2 (**Figures 1 and S7)**. Negative associations between baseline GlycA and entorhinal cortex volume was observed for the follow-up years 2 (*p*=0.011, ρ=-0.140), 4 (*p*=0.040, ρ=-0.165), 6 (*p*=0.024, ρ=-0.224), 7 (*p*=0.026, ρ=-0.257) and 8 (*p*=0.025, ρ=-0.345) (**Figure 4**). The negative correlation between longitudinal entorhinal cortex volume and baseline GlycA in participants diagnosed with LMCI at baseline was confirmed using a linear mixed model (**Table S3**). The LMCI-GlycA interaction to predict entorhinal cortex volume was confirmed using a linear mixed model (*p*=0.005, **Table S1**). Additionally, GlycA-entorhinal cortex volume association among participants with LMCI was not-sex dependent (*p*_interaction_=0.333). Similar associations were not observed with other brain regional volumes (**Table S4**).

**Figure 4.**
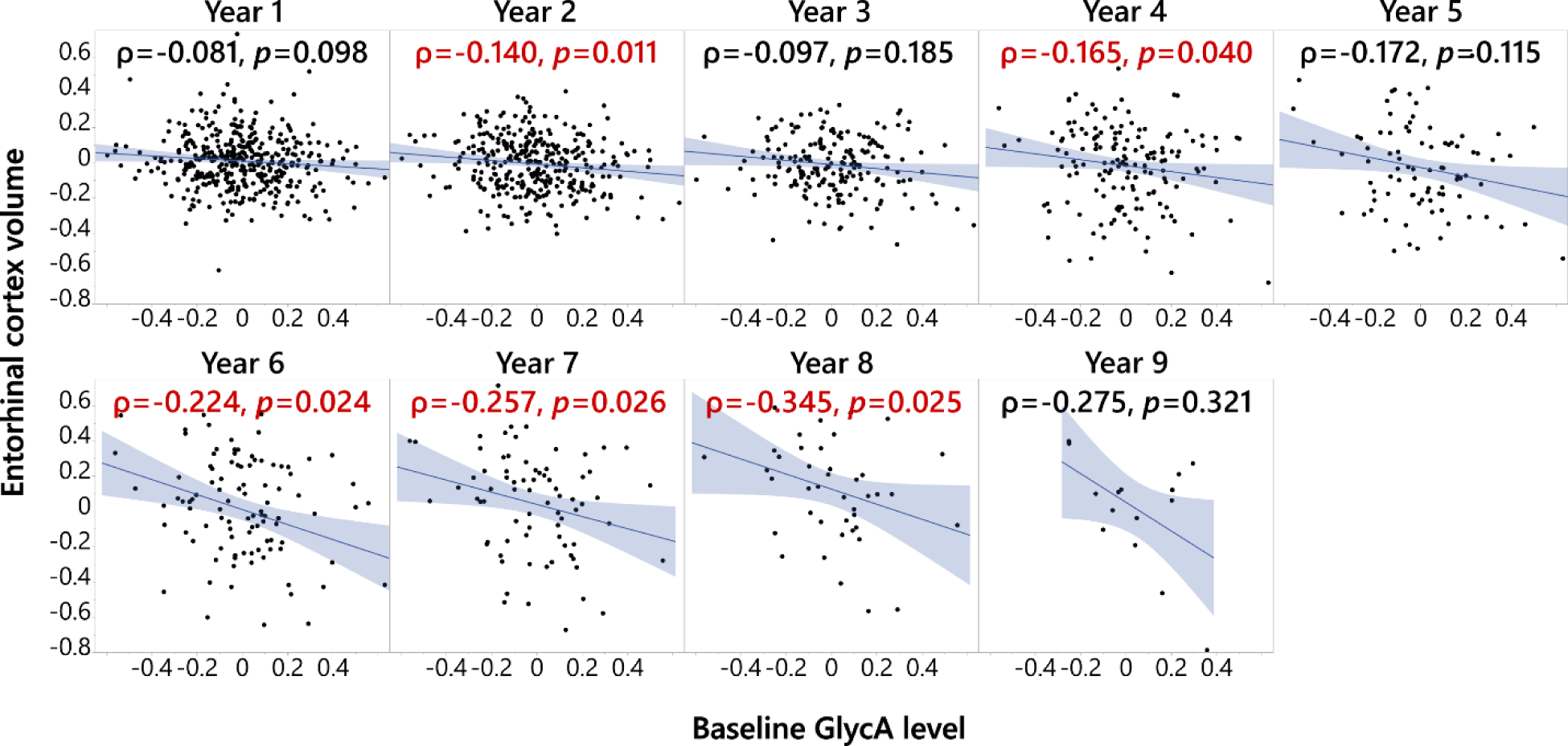
Baseline GlycA level was associated with entorhinal cortex volume in late mild cognitive impairment (LMCI) participants at the 2^nd^, 4^th^, and 6-8^th^ years of follow-up. Spearman’s rank order correlation was performed on residuals of entorhinal cortex volume and GlycA, and the analyses were stratified by diagnosis status. Entorhinal cortex volume at different years was adjusted for baseline entorhinal cortex volume, screening age, follow-up year, BMI at the time, APOE4, sex, and education, treating participant ID as random factors (all MRI volumes were adjusted for intracranial volume and magnet type); baseline GlycA level was adjusted for medication, screening age, baseline BMI, APOE, sex, and education. The analysis was performed on participants with different diagnosis status, but only in the participants with LMCI was entorhinal cortex volume in continuous follow-up years negatively associated with baseline GlycA level. Therefore, the analysis results for the participants with LMCI are shown here. In addition, the LMCI-GlycA interaction was confirmed with a linear mixed model (**Table S1**). A similar full factorial linear mixed model was used to show that there were no sex-GlycA interactions controlling DX (*p*=0.564). *Abbreviations:* BMI: Body mass index; DX: Diagnosis at baseline; GlycA: Glycoprotein acetyls; ID: Identification; MRI: Magnetic resonance imaging.

### Baseline GlycA Correlated with Baseline Brain Structural Atrophies Specifically in Female Participants Diagnosed with LMCI and AD

To identify the cross-sectional relationship between GlycA and regional brain atrophies measured by MRI, we used Spearman’s rank order correlation with sex and diagnostic group stratification (**Table S5, Figure 5**). Bilateral total/mean measurements were used to examine the correlation of brain atrophy with GlycA, with measures in the left and right hemispheres showing similar results (**Table S6**). In addition, rather than using thickness, we mainly used regional volumes to describe brain atrophy due to its stronger sensitivity (51), though we did provide both results (**Table S6**). Comparisons of volumetric measurements among participants of different diagnosis status were performed using an ANCOVA model (**Table S7**).

**Figure 5.**
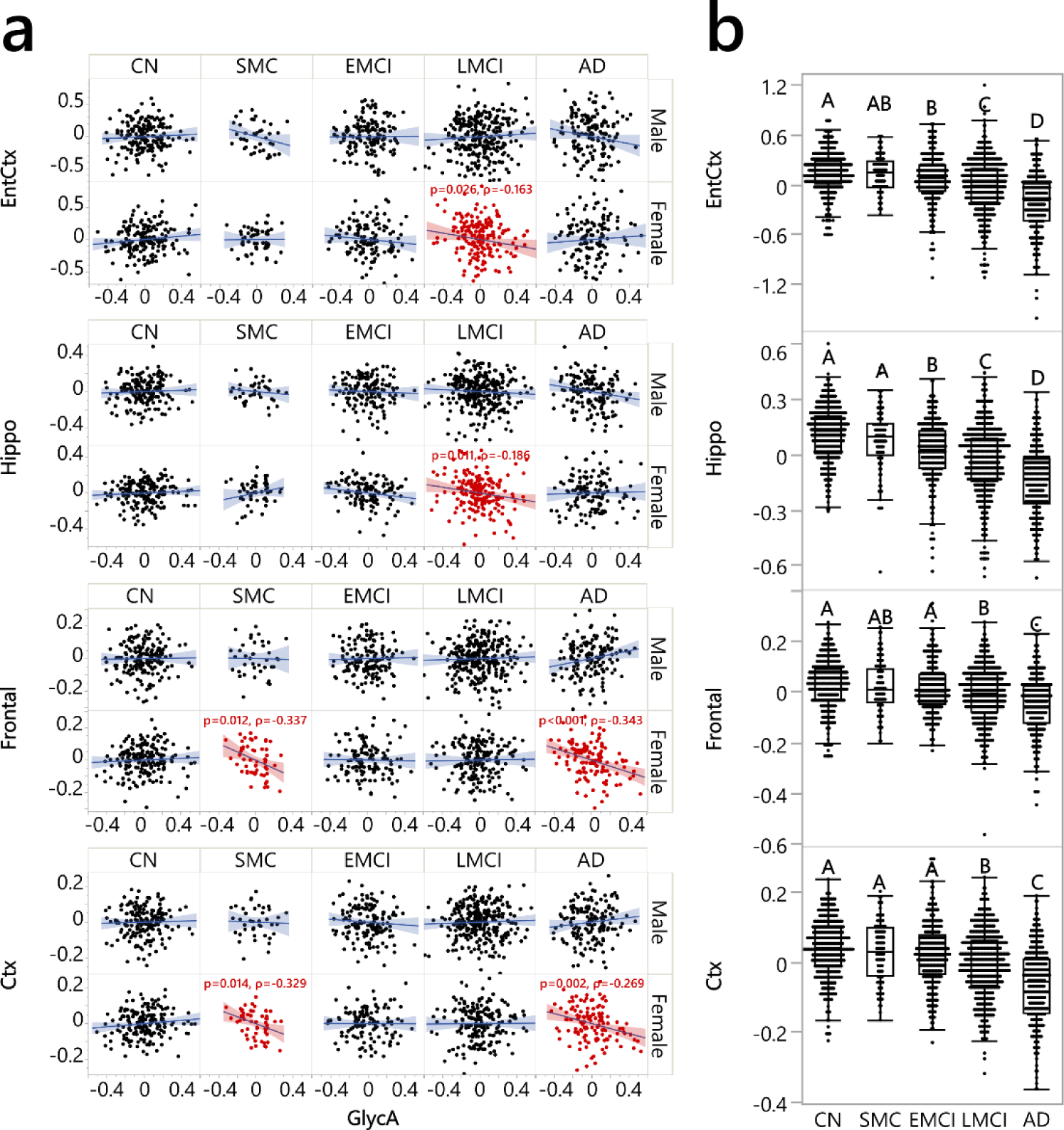
GlycA was associated with regional brain volumes in females with SMC, LMCI and AD. The full results are listed in **Tables S2 and S3** and the present figure illustrates selected representative examples. **a)** Significant associations were found between GlycA and volumetric measurements of brain regions, i.e., entorhinal cortex (EntCtx), hippocampus (Hippo), frontal lobe (Frontal), and cerebral cortex grey matter (Ctx). Spearman’s rank order correlation was performed between the residual of GlycA (log2-transformed, adjusted for medications, APOE4, age, BMI, and education) and residuals of whole brain regional volumes (log2-transformed, adjusted for log2-intracranial volume, magnet strength/scan type, APOE4, age, BMI, education); the analysis was performed stratified by sex and diagnosis groups. **b)** This panel indicates different levels of these brain volumetric measurements, adjusting for the same confounders without stratifications (**Table S7**). There were no significant sex-diagnosis stages interactions (*p*_interaction_=0.284 (EntCtx), 0.244 (Hippo), 0.324 (Frontal) and 0.398 (Ctx)) for brain volumetric measurements and therefore this panel is not divided by sex. N: for males: CN: 178, SMC: 40, EMCI: 154, LMCI: 292, and AD: 154; for females: CN: 183, SMC: 55, EMCI: 125, LCMI: 187, and AD: 13. *Abbreviations:* AD: Alzheimer’s disease; ANCOVA: Analysis of covariance; BMI: Body mass index; CN: Cognitively normal; EMCI: Early mild cognitive impairment; GlycA: Glycoprotein acetyls; LMCI: Late mild cognitive impairment; SMC: Significant memory concerns.

In females with AD, GlycA was negatively associated with the frontal lobe volume (*p*<0.001, ρ=-0.343), cerebral cortex grey matter volume (*p*=0.002, ρ=-0.269), global grey matter mean volume (*p*=0.002, ρ=-0.267), sensory motor volume (*p*=0.014, ρ=-0.214), and parietal lobe volume (*p*=0.011, ρ=-0.222) (**Table S5**). Except for parietal lobe volume, all of these GlycA-brain morphology associations were specific for females with AD, compared to males with AD. The specificity of the association was measured using a linear factorial model with GlycA-sex interaction, stratified by diagnostic group, with results for AD participants including *p_interaction_* <0.001 for the frontal lobe volume, *p_interaction_* =0.008 for cerebral cortex grey matter volume, *p_interaction_* =0.007 for grey matter mean volume, and *p_interaction_* =0.020 for sensory motor volume (**Table S5**).

In addition, in females with LMCI, GlycA was negatively associated with hippocampus volume (*p*=0.011, ρ=-0.186) and entorhinal cortex volume (*p*=0.026, ρ=-0.163). In females with SMC, GlycA was negatively associated with cerebral cortex grey matter volume (*p*=0.014, ρ=-0.329), frontal lobe volume (*p*=0.012, ρ=-0.337), and mean grey matter volume (*p*=0.011, ρ=-0.341). No correlation passing false discovery rate correction was observed in the CN or EMCI diagnostic groups, nor in male participants of any diagnosis status. None of the GlycA-sex interactions in non-AD groups and GlycA-diagnosis interactions in both sexes were significant after false discovery rate correction (**Table S5**).

It is important to note that brain region atrophy was not uniform along AD disease progression (**Figure 5b, Table S7**). While areas such as the entorhinal cortex and hippocampus had significantly decreased volumes compared to CN as early as the EMCI stage (*p*<0.05), other volumetric measurements – such as those in cerebral cortex grey matter and the frontal lobe – only started to have significantly decreased volumes compared to CN in LMCI. Consistent with this, the correlations between GlycA and hippocampus volume and between GlycA and entorhinal cortex volume were found in an earlier diagnosis stage in females (i.e., LMCI) compared to GlycA correlation with cerebral cortex grey matter volume and frontal lobe volume, which were found in a later stage (i.e., AD). However, such disease state patterns were not inclusive, meaning that not all brain anatomical region changes along disease progression coincided with their correlation with GlycA. An example of this is the early change of medial temporal lobe volume at the EMCI stage without a significant correlation with GlycA, found in participants with any diagnosis status.

### Association of Plasma GlycA with CSF A/T/N Biomarkers

We investigated the relationship between GlycA in plasma and A/T/N biomarkers in CSF using Spearman’s rank correlation as described above. No association was observed between plasma GlycA and A/T/N biomarkers in CSF, including the biomarkers Aβ 42, pTau/Aβ 42, pTau, total Tau, and total Tau/Aβ 42 (**Table S8**). GlycA in plasma was also not associated with A/T/N biomarkers in CSF in all participants in the continuous future follow-up years (**Table S9**).

## DISCUSSION

The current study describes associations between the level of peripheral inflammation and AD-related structural and cognitive changes in the CNS. Importantly, these peripheral-central correlations manifest primarily in the later stages of the progression toward AD, which emphasizes the diagnosis-status-specific heterogeneity in the relationship between peripheral inflammation, cognition, and brain atrophy. Our findings point to peripheral inflammation as a risk factor for future cognitive decline and brain atrophy in both males and females, which opens the door to the identification of a population at risk as well as potential therapeutic intervention.

Abnormal cognition and memory decline are important markers of AD pathogenesis (52), and the structural and functional degeneration of the entorhinal cortex often indicate the early changes in AD development, which implies the particular vulnerability of this region to AD pathogenesis (53, 54). Accordingly, our results have shown that the peripheral inflammation at baseline correlates with decreasing executive functions, decreasing memory and decreasing entorhinal cortex volume in the follow-up years. The decreased entorhinal cortex volume may provide an inflammatory-related structural basis to explain the observed declines in brain functions that are associated with inflammation.

In addition, such observations between baseline GlycA and future declines in executive functions, memory and entorhinal cortex volume are restricted to the participants diagnosed with LMCI at baseline. Among non-AD participants, the LMCI diagnosis group had the earliest brain function and structural decline in the follow-up years. Mechanistically, peripheral cytokines can directly enter the CNS and/or interact with the blood-brain barrier through multiple pathways (13, 55). Meanwhile, under pathological conditions, peripheral immune cells can migrate across the blood-brain barrier (56–58) and modulate microglia function (59, 60), a key player in CNS inflammation and AD development. As a result, peripheral inflammation stimuli can modulate the expression of inflammation-related genes in the brain (19) and exacerbate CNS neurodegeneration (53). Brain inflammation has been suggested to have differential effects along the AD progression (20). For example, in the AD rodent model of APPPS1 mice, the lack of neuroinflammation-related gene TREM2 expression (i.e., TREM2 deficiency) lowered amyloid pathologies at the earlier stage of disease development but worsened it at the later stage.

Specifically at the later stage, TREM2 deficiency reduced the proliferation of myeloid cells and the gene transcription levels related to inflammation, such as IL-1β and TNF-α (20). These studies support the notion that certain inflammation components can be protective at the beginning of the progression toward AD, but detrimental to CNS homeostasis at the later stages (20, 61). Such a notion not only explains the relationship between baseline GlycA and AD-related biomarkers in the follow-up years for participants with LMCI, but also the cross-sectional correlation between baseline GlycA and baseline AD-related biomarkers in the female participants with LMCI and AD.

Sex differences were observed in the cross-sectional correlation between peripheral inflammation and brain structures. This was different from the relationship between baseline peripheral inflammation and brain structures in the follow-up years, in which no sex-dependent effect was observed. While causation cannot be inferred from our observations, such a difference on peripheral-central connection between cross-sectional and longitudinal analysis may be due to the difference between acute and chronic inflammation, both of which can be captured by GlycA (26, 29). Sexual dimorphism in neurological disorders has been associated with differential microglia sub-populations, including disease-associated microglia and activated response microglia (62–64), as well as the microglial involvement in AD-related pathologies such as neuroinflammation (65), Aβ accumulation (62) and tau pathologies (65–67). In a rodent study, the identical acute peripheral inflammatory stimuli caused a more substantial pro-inflammatory response in the aged brains of females compared to males (18). However, it is not clear whether the aged brains of males and females respond to the chronic peripheral stimuli in a similar manner (19, 68), considering that males and females overall manifest different fluctuations in peripheral inflammation over a lifetime (69). Further longitudinal studies on aged models or populations with chronic inflammation are therefore warranted to clarify the potential sex effect of inflammation on AD development.

Additionally, while we observed the correlation between GlycA and brain structure, and between GlycA and cognitive changes, there was no correlation between GlycA and CSF biomarkers for Aβ and tau pathologies. Therefore, additional regulation of peripheral inflammation and AD pathologies must occur in CNS. Support for this notion can be found in animal research. For example, amyloid precursor protein is negatively associated with CSF Aβ level (70). However, in an amyloid precursor protein knock-in mouse model, peripheral inflammation increased Aβ deposition in the brain (71), but interrupted Aβ clearance from CSF to blood (71). Such an interaction could explain the lack of changes in CSF Aβ under the peripheral inflammation stimuli (71), consistent with our current observation. In addition, peripheral inflammation affects the transmission of Tau proteins among brain regions (72), but how these processes collectively regulate the level of Tau pathology in CSF is not clear. Overall, the mechanisms that regulate the complex crosstalk between peripheral inflammation and different AD pathologies warrant further investigation.

In conclusion, our study describes connections between peripheral inflammation and cognitive and structural AD-related changes in the brain. Moreover, it highlights the implication of systemic inflammation at earlier timepoints on the later stage toward AD and highlights the potentially higher sensitivity or susceptibility of females, but not males, to systemic inflammation. These findings contribute to our understanding of disease heterogeneity and lay the groundwork for more personalized approaches toward disease treatment.

## LIMITATIONS

It should be pointed out that our analyses were limited to observational findings instead of causative ones. Replications on independent cohorts are required for confirmation. In addition, the comorbidity of AD pathologies and the diagnosis status of inflammatory diseases such as diabetes and metabolic syndrome were not discussed here, with the consideration of medication-adjusted GlycA as an overall peripheral inflammatory biomarker without knowing such diagnosis statuses. The comorbidity of diabetes and AD-related pathologies has been reported in a previous study performed using the ADNI data (73).

## Data Availability

All data were acquired from the ADNI database (www.adni-info.org), managed through the Laboratory of Neuro Imaging Image & Data Archive (http://adni.loni.usc.edu/).

## ACKNOWLEDGEMENTS

Dr. Kamil Borkowski is funded by the National Institutes of Health grant P30AG072972. Dr. Kwangsik Nho is funded by grants U01 AG072177, R01 LM012535, and U19AG074879. The Alzheimer’s Disease Metabolomics Consortium (ADMC) is funded wholly or in part by the following National Institute on Aging grants and supplements, which are components of the Accelerating Medicines Partnership for AD (AMP-AD) and/or Molecular Mechanisms of the Vascular Etiology of AD (M2OVE-AD): NIA R01AG046171, RF1AG051550, RF1AG057452, R01AG059093, RF1AG058942, U01AG061359, U19AG063744, 3U19AG063744-04S1, 1R01AG081322, and FNIH: #DAOU16AMPA awarded to Dr. Rima Kaddurah-Daouk at Duke University in partnership with a large number of academic institutions. Dr. John W. Newman was supported by USDA Project 2032-51530-025-00D. The USDA is an equal opportunity provider and employer. Dr. Peter J. Meikle is supported by an Investigator grant (2009965) from the National Health and Medical Research Council of Australia. This work was supported by the Victorian Government’s Operational Infrastructure Support Program.

Data collection and sharing for this project was supported by the Alzheimer’s Disease Neuroimaging Initiative (ADNI) with National Institutes of Health Grant U01 AG024904 and DOD ADNI with Department of Defense award number W81XWH-12-2-0012. ADNI is funded by the following entities: the National Institute on Aging, the National Institute of Biomedical Imaging and Bioengineering, and through generous contributions from the following: AbbVie, Alzheimer’s Association; Alzheimer’s Drug Discovery Foundation; Araclon Biotech; BioClinica, Inc.; Biogen; Bristol-Myers Squibb Company; CereSpir, Inc.; Cogstate; Eisai Inc.; Elan Pharmaceuticals, Inc.; Eli Lilly and Company; EuroImmun; F. Hoffmann-La Roche Ltd. and its affiliated company Genentech, Inc.; Fujirebio; GE Healthcare; IXICO Ltd.; Janssen Alzheimer Immunotherapy Research & Development, LLC.; Johnson & Johnson Pharmaceutical Research & Development LLC.; Lumosity; Lundbeck; Merck & Co., Inc.; Meso Scale Diagnostics, LLC.; NeuroRx Research; Neurotrack Technologies; Novartis Pharmaceuticals Corporation; Pfizer Inc.; Piramal Imaging; Servier; Takeda Pharmaceutical Company; and Transition Therapeutics. ADNI clinical sites in Canada were supported by the Canadian Institutes of Health Research. Private sector contributions are facilitated by the Foundation for the National Institutes of Health (www.fnih.org). The grantee organization is the Northern California Institute for Research and Education, and the study is coordinated by the Alzheimer’s Therapeutic Research Institute at the University of Southern California. ADNI data are disseminated by the Laboratory for Neuro Imaging at the University of Southern California.

The editorial services of Mr. Jon Kilner, MS, MA (Pittsburgh) and the assistance of Ms. Lisa Howerton (Duke) are acknowledged here.

