## Supplemental material for "Peripheral inflammation is associated with structural brain atrophy and cognitive decline linked to mild cognitive impairment and Alzheimer’s disease"

### SUPPLEMENTARY MATERIALS

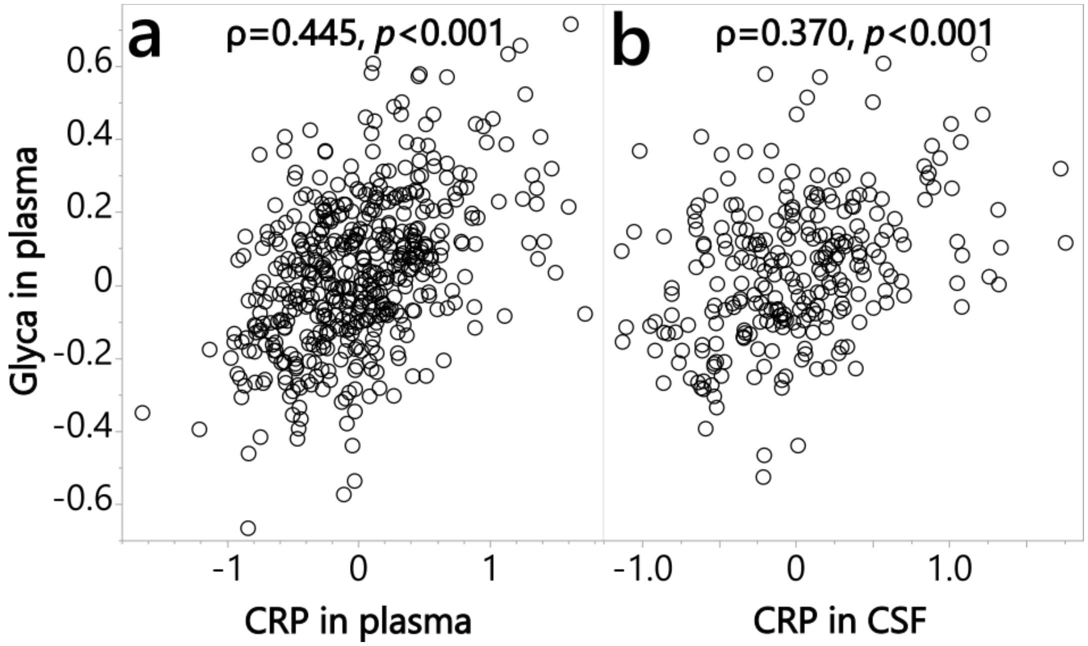

**Supplementary Figure S1. Plasma GlycA was associated with a common inflammatory marker, CRP, in both plasma and CSF.**
Spearman correlation between **a)** plasma GlycA and CRP in plasma (n=484) and **b)** plasma GlycA and CSF (n=267). All data were normalized and medication-adjusted; participants' diagnosis status ranged from healthy controls to patients with Alzheimer's disease. *Abbreviations:* CRP: C-reactive protein; CSF: Cerebrospinal fluid; GlycA: Glycoprotein acetyls.

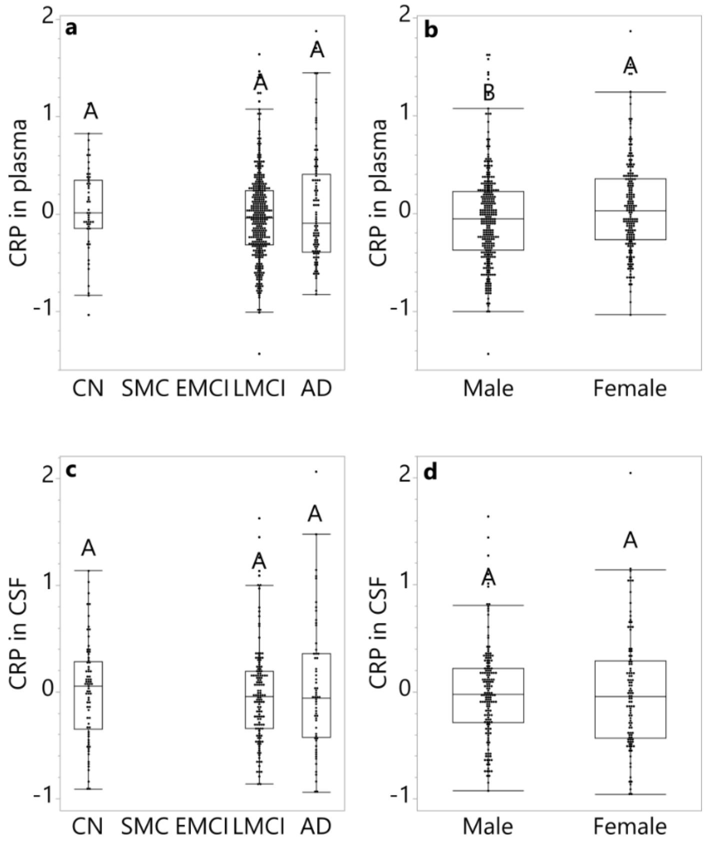

**Supplementary Figure S2. CRP in CSF and Blood.**
CRP levels were measured in plasma, with participants **a)** grouped by diagnosis status, log2-transformed and adjusted for medications, sex, age, APOE4, BMI, and education level, and **b)** grouped by sex, log2-transformed and adjusted for medications, diagnosis status, age, APOE4, BMI, and education level. CRP levels were measured in CSF, with participants **c)** grouped by diagnosis status, log2-transformed and adjusted for medications, sex, age, APOE4, BMI, and education level, and **d)** grouped by sex, log2-transformed and adjusted for medications, diagnosis status, age, APOE4, BMI, and education level. ANCOVA and Tukey HSD post hoc analysis were performed; levels not labelled with the same letter differed significantly ($p$ <0.05). Values in the bar graph were shown as the mean residual CRP level ± standard error.
*Abbreviations:* AD: Alzheimer's disease; CN: Cognitively normal; CRP: C-reactive protein; CSF: cerebrospinal fluid; EMCI: Early mild cognitive impairment; LMCI: Late mild cognitive impairment; SMC: Significant memory concerns.

| Year follow-up | 1 | 2 | 3 | 4 | 5 | 6 | 7 | 8 | 9 | 10 | 11 | 12 | 13 |
|---|---|---|---|---|---|---|---|---|---|---|---|---|---|
| CN | A | A | AB | AB | AB | BC | CD | DE | EF | F | DEF | EF | F |
| SMC | A | A | | A | AB | B | | | | | | | |
| EMCI | A | A | AB | A | BC | ABC | C | C | BC | | | | |
| LMCI | A | A | B | C | CD | DE | DE | DE | E | E | | | |
| AD | A | B | | | | | | | | | | | |

* Heatmap scale:

| | | |
|---|---|---|
| 1.05 | 0.00 | -1.51 |

**Supplementary Figure S3. Executive function (EF) progression heatmap in participants diagnosed with CN, SMC, EMCI, LCMI and AD at baseline.**
Color scale indicates the least square mean, while the letters annotate the Tukey/t-test analysis results in each diagnose status group, in an ANCOVA model with covariant of year of follow-up, baseline EF, age at screening, BMI at visit, sex, APOE4, and education level; participant ID as a random factor; and stratified by baseline diagnosis. Data points with less than 10 participants were removed; the months of 6 and 18 were not used to simplify illustration. Results here indicates that the EF of participants with LMCI and AD experienced decline faster than participants with CN, SMC, and EMCI.
*Abbreviations:* AD: Alzheimer's disease; BMI: Body mass index; CN: Cognitively normal; EMCI: Early mild cognitive impairment; LMCI: Late mild cognitive impairment; SMC: Significant memory concerns.

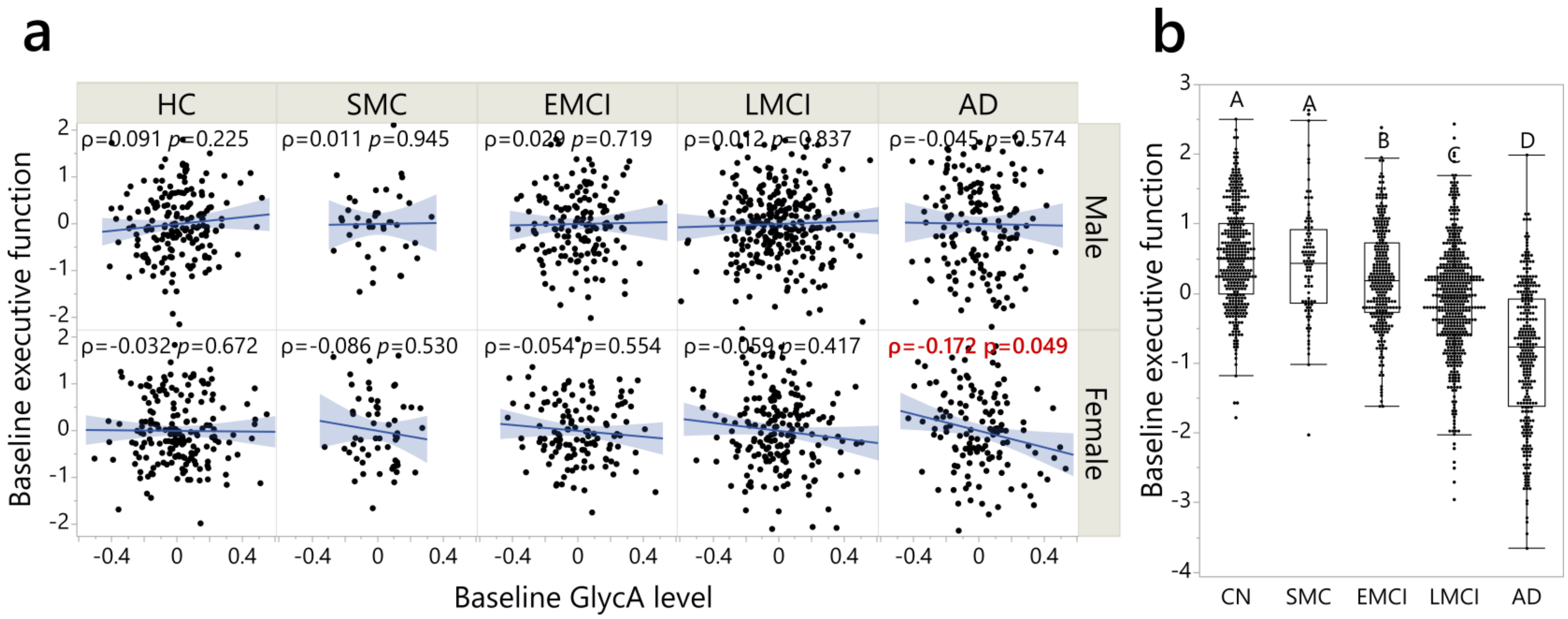

**Supplementary Figure S4. GlycA was cross-sectionally associated with brain executive functioning (EF) in females with AD.**
**a)** Spearman's rank order correlation results, which was performed between the residual of GlycA (log2-transformed, adjusted medications, APOE4, age, BMI, and education) and executive functioning composite score (adjusted APOE4, age, BMI, and education); all analyses were performed stratified by sex and diagnosis groups. Even though there was no GlycA-sex interaction for EF at different diagnosis status, the sex and diagnosis status stratifications were applied due to the high GlycA level in female participants and in participants with AD (**Figure 2**). **b)** Different levels of EF by diagnosis status, controlling for APOE4, age, BMI, education, and sex. There were no significant sex-diagnosis stages interaction ($p$=0.717) for EF.
*Abbreviations:* AD: Alzheimer's disease; BMI: Body mass index; CN: Cognitively normal; EMCI: Early mild cognitive impairment; GlycA: Glycoprotein acetyls; LMCI: Late mild cognitive impairment; SMC: Significant memory concerns.

**a)**

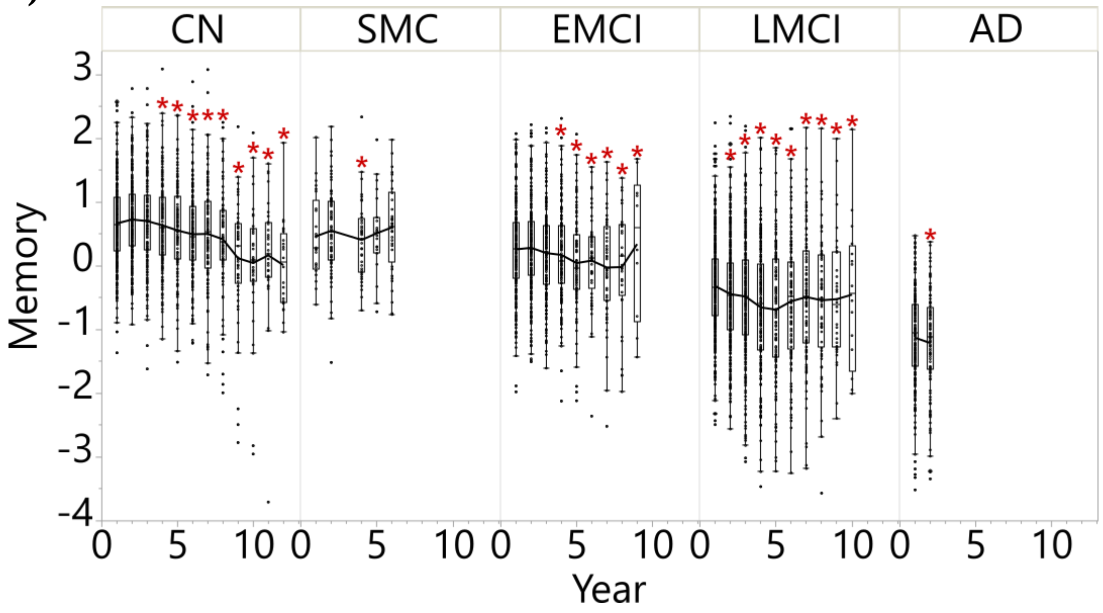

**b)**

| Year follow up | 1 | 2 | 3 | 4 | 5 | 6 | 7 | 8 | 9 | 10 | 11 | 12 |
|---|---|---|---|---|---|---|---|---|---|---|---|---|
| CN | AB | A | AB | BC | CD | DE | DE | EF | FG | G | FG | G |
| SMC | A | A | | B | AB | AB | | | | | | |
| EMCI | A | A | AB | BC | CD | BCD | D | CD | CD | | | |
| LMCI | A | B | C | D | DE | DEF | EFG | FGH | GH | H | | |
| AD | A | B | | | | | | | | | | |

* Heatmap scale:

| | | |
|---|---|---|
| 1.17 | 0.00 | -1.28 |

**Supplementary Figure S5. Memory (MEM) progression in participants diagnosed with CN, SMC, EMCI, LCMI and AD at baseline.**
**a)** Memory was memory composite score adjusted by sex, ApoE4, education, BMI at visit and age at screening. Results annotated with * in a) were at a significantly different level compared to Year 1 within the diagnosis group as indicated in b). **b)** The color scale indicates the least square mean, while the letters annotate the Tukey/t-test analysis results in each diagnose status group, in an ANCOVA model with a covariant of year of follow-up, baseline MEM, age at screening, BMI at visit, sex, APOE4, and education level; participant ID as a random factor; and stratified by baseline diagnosis. Data points with less than 10 participants were removed; the months of 6 and 18 were not used to simplify illustration. Results here indicated that the MEM of participants with LMCI and AD experienced decline faster than participants with CN, SMC, and EMCI.
*Abbreviations:* AD: Alzheimer's disease; ANCOVA: Analysis of covariance; BMI: Body mass index; CN: Cognitively normal; EMCI: Early mild cognitive impairment; LMCI: Late mild cognitive impairment; SMC: Significant memory concerns.

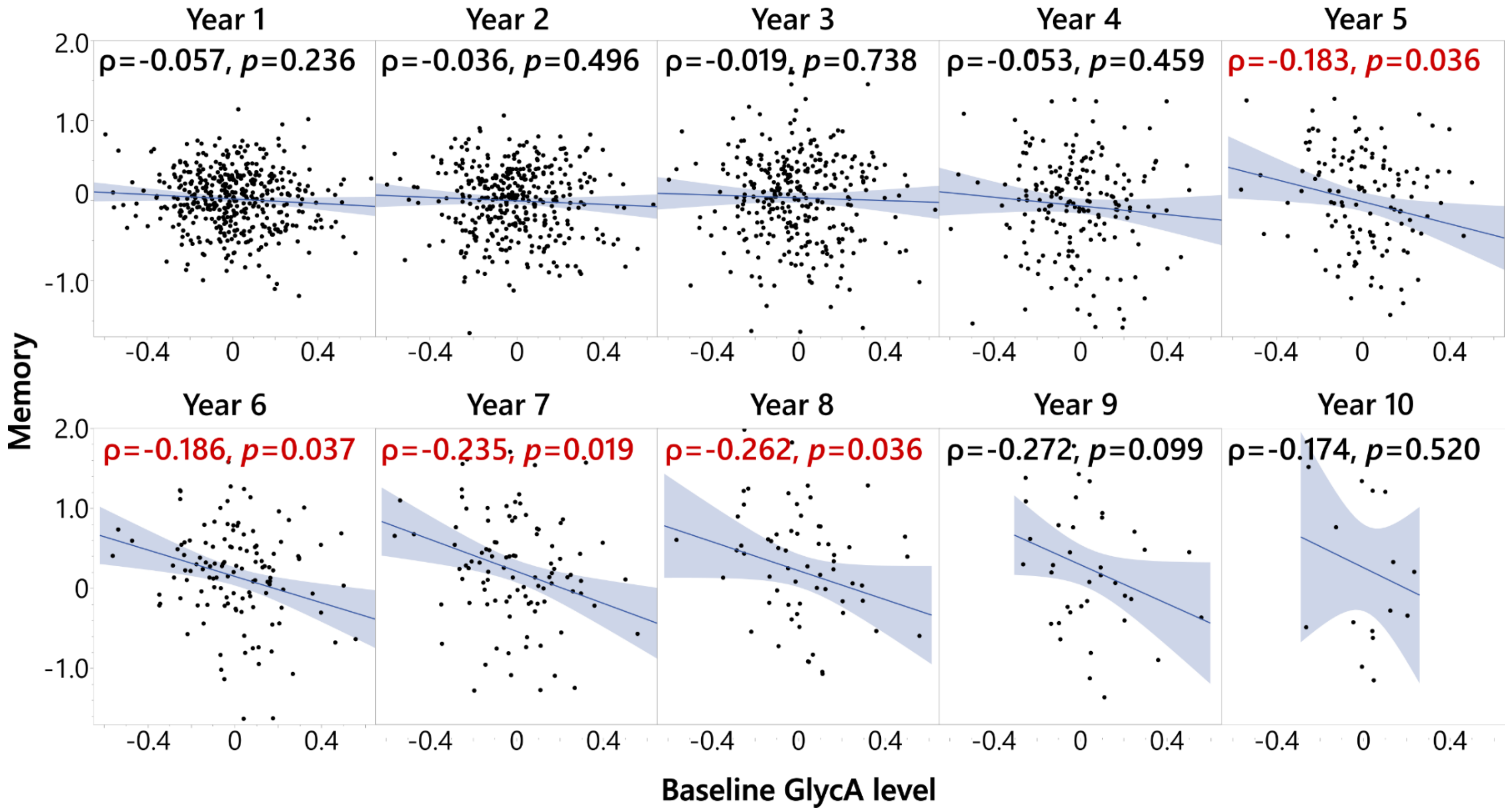

**Supplementary Figure S6. Baseline GlycA level was associated with memory composite score (MEM) in late mild cognitive impairment (LMCI) participants at 5-8 years follow-up.**

Spearman's rank order correlation was performed on residuals of MEM and GlycA, and the analyses were stratified by diagnosis status. MEM at different years were adjusted for baseline MEM level, screening age, follow up year, BMI at the time, APOE4, sex, and education, treating participants' ID as random factors; baseline GlycA levels were adjusted for medication, screening age, baseline BMI, APOE, sex, and education. The analysis was performed on participants with different diagnosis status, but only in the participants with LMCI was MEM at continuous follow-up years negatively associated with baseline GlycA level. Therefore, the analysis results for the participants with LMCI is shown here. A full factorial linear mixed model was used to shown that there were no DX-GlycA interactions in LMCI ($p$ =0.441, **Table S1**) nor sex-GlycA interactions controlling DX ($p$=0.864).

*Abbreviations:* BMI: Body mass index; DX: Diagnosis at baseline; EMCI: Early mild cognitive impairment; GlycA: Glycoprotein acetyls; ID: Identification.

| Year | 1 | 2 | 3 | 4 | 5 | 6 | 7 | 8 | 9 | 10 | 11 | 12 | 13 |
|---|---|---|---|---|---|---|---|---|---|---|---|---|---|
| CN | A | AB | AB | BC | BCD | CDE | FG | DEF | EFG | EFGH | EFGH | GH | H |
| SMC | AB | A | | A | AB | B | | | | | | | |
| EMCI | A | AB | BC | BC | ABC | C | C | C | ABC | | | | |
| LMCI | A | B | C | D | DE | E | E | DE | F | | | | |
| AD | A | B | | | | | | | | | | | |

* Heatmap scale:

| 0.17 | 0.00 | -0.47 |
|---|---|---|

**Supplementary Figure S7. Entorhinal cortex (EC) volume progression heatmap in participants diagnosed with CN, SMC, EMCI, LCMI and AD.**

The color scale indicates the least square mean, while the letters annotate the Tukey/t-test analysis results in each diagnose status groups, in an ANCOVA model with covariant of year of follow-up, baseline EC volume, screening age, sex, APOE4, BMI at visit and education level; and participant ID as a random factor. All EC volume are log2-transformed and adjusted for magnet type and log2-ICV. Data points with less than 10 participants were removed; the months of 3, 6 and 18 were not used to simplify illustration. Results here indicate that the EC volume of participants with LMCI and AD experienced atrophy faster than participants with CN, SMC, and EMCI.

*Abbreviations:* AD: Alzheimer's disease; ANCOVA: Analysis of covariance; BMI: Body mass index; CN: Cognitively normal; EMCI: Early mild cognitive impairment; ICV: intracranial volume; LMCI: Late mild cognitive impairment; SMC: Significant memory concerns.

**Supplementary Table S1. Linear mixed model to assess the interaction between baseline GlycA and diagnosis status for predicting longitudinal EF/MEM/EntCtx, adjusting age, BMI, baseline EF/MEM/EntCtx, month follow-up, sex, ApoE4, and education, treating subject ID as random effect.**

**Parameters Estimates**

| **Variables: EF [1]** | Estimate | Std Error | DFDen | t Ratio | Prob>\|t\| |
|---|---|---|---|---|---|
| Intercept | 0.164454 | 0.099066 | 1394 | 1.66 | 0.097 |
| DX[CN]*(GlycA[2]+0.00563) | 0.211384 | 0.174711 | 1356 | 1.21 | 0.227 |
| DX[SMC]*(GlycA[2]+0.00563) | 0.01732 | 0.371251 | 1547 | 0.05 | 0.963 |
| DX[EMCI]*(GlycA[2]+0.00563) | 0.188913 | 0.193427 | 1370 | 0.98 | 0.329 |
| DX[LMCI]*(GlycA[2]+0.00563) | -0.35729 | 0.158973 | 1425 | -2.25 | 0.025 |
| DX[CN] | 0.239153 | 0.032233 | 1349 | 7.42 | <0.001 |
| DX[SMC] | 0.182289 | 0.057984 | 1521 | 3.14 | 0.002 |
| DX[EMCI] | 0.115754 | 0.034808 | 1352 | 3.33 | 0.001 |
| DX[LMCI] | -0.15716 | 0.02998 | 1403 | -5.24 | <0.001 |
| GlycA[2] | -0.14775 | 0.119449 | 1526 | -1.24 | 0.216 |
| Sex | 0.025016 | 0.01703 | 1367 | 1.47 | 0.142 |
| ApoE4 | 0.081882 | 0.01716 | 1370 | 4.77 | <0.001 |
| Education | 0.002876 | 0.006061 | 1366 | 0.47 | 0.635 |
| Year follow up | -0.08406 | 0.003331 | 4264 | -25.24 | <0.001 |

[1]: adjusted age and BMI at the time and baseline EF;
[2]: adjusted medication, screening age and baseline BMI

| Random Effect | Var Ratio | Var Component | Std Error | 95% Lower | 95% Upper | Wald p-Value | % of Total |
|---|---|---|---|---|---|---|---|
| Participant ID | 1.095423 | 0.268384 | 0.01344 | 0.242042 | 0.294725 | <.0001 | 52.277 |
| Residual | | 0.245005 | 0.005616 | 0.234359 | 0.256396 | | 47.723 |
| Total | | 0.513388 | 0.013991 | 0.487034 | 0.541952 | | 100 |

| **Variables: MEM[1]** | Estimate | Std Error | DFDen | t Ratio | Prob>\|t\| |
|---|---|---|---|---|---|
| Intercept | 0.205727 | 0.070564 | 1414 | 2.92 | 0.004 |
| DX[CN]*(GlycA[2]+0.00564) | 0.090745 | 0.124461 | 1372 | 0.73 | 0.466 |
| DX[SMC]*(GlycA[2]+0.00564) | 0.025396 | 0.263563 | 1565 | 0.1 | 0.923 |
| DX[EMCI]*(GlycA[2]+0.00564) | -0.03293 | 0.137831 | 1385 | -0.24 | 0.811 |
| DX[LMCI]*(GlycA[2]+0.00564) | -0.08712 | 0.113133 | 1436 | -0.77 | 0.441 |
| DX[CN] | 0.081501 | 0.02297 | 1367 | 3.55 | 0.000 |
| DX[SMC] | 0.016512 | 0.041259 | 1540 | 0.4 | 0.689 |
| DX[EMCI] | 0.046266 | 0.024813 | 1368 | 1.86 | 0.063 |
| DX[LMCI] | -0.06633 | 0.021368 | 1419 | -3.1 | 0.002 |
| GlycA[2] | -0.12042 | 0.084895 | 1541 | -1.42 | 0.156 |
| Sex | 0.027531 | 0.012148 | 1386 | 2.27 | 0.024 |
| ApoE4 | 0.081963 | 0.012229 | 1388 | 6.7 | 0.000 |
| Education | -0.00043 | 0.004316 | 1385 | -0.1 | 0.922 |

| | | | | | |
|---|---|---|---|---|---|
| Year follow up | -0.07109 | 0.002449 | 4327 | -29.03 | 0.000 |

[1]: adjusted age and BMI at the time and baseline MEM
[2]: adjutsed medictaion, screening age and baseline BMI

| Random Effect | Var Ratio | Var Component | Std Error | 95% Lower | 95% Upper | Wald p-Value | % of Total |
|---|---|---|---|---|---|---|---|
| Participant ID | 1.054251 | 0.135832 | 0.006793 | 0.122519 | 0.149145 | <.0001 | 51.32 |
| Residual | | 0.128842 | 0.002933 | 0.123282 | 0.13479 | | 48.68 |
| Total | | 0.264674 | 0.007111 | 0.251272 | 0.279183 | | 100 |

| **Variables: EntCtx[1]** | Estimate | Std Error | DFDen | t Ratio | Prob>\|t\| |
|---|---|---|---|---|---|
| Intercept | 0.075462 | 0.024712 | 1345 | 3.05 | 0.002 |
| DX[CN]*(GlycA[2]+0.00516) | -0.03511 | 0.043557 | 1284 | -0.81 | 0.42 |
| DX[SMC]*(GlycA[2]+0.00516) | 0.127813 | 0.096816 | 1572 | 1.32 | 0.187 |
| DX[EMCI]*(GlycA[2]+0.00516) | 0.000871 | 0.048991 | 1393 | 0.02 | 0.986 |
| DX[LMCI]*(GlycA[2]+0.00516) | -0.11209 | 0.040149 | 1381 | -2.79 | 0.005 |
| DX[CN] | 0.032834 | 0.008017 | 1289 | 4.1 | <0.001 |
| DX[SMC] | 0.041378 | 0.014818 | 1557 | 2.79 | 0.005 |
| DX[EMCI] | 0.018222 | 0.008768 | 1356 | 2.08 | 0.038 |
| DX[LMCI] | -0.03359 | 0.007539 | 1384 | -4.46 | <0.001 |
| GlycA[2] | -0.03025 | 0.03072 | 1533 | -0.98 | 0.325 |
| Sex | 0.014059 | 0.004262 | 1312 | 3.3 | 0.001 |
| ApoE4 | 0.015549 | 0.004293 | 1322 | 3.62 | <0.001 |
| Education | -0.00014 | 0.001507 | 1301 | -0.09 | 0.928 |
| Year follow up | -0.02405 | 0.001037 | 3579 | -23.18 | <0.001 |

[1]: adjusted age and BMI at the time and baseline EntCtx, after adjusting all log2-volume with log2-ICV and magnet type
[2]: adjusted medication, screening age and baseline BMI

| Random Effect | Var Ratio | Var Component | Std Error | 95% Lower | 95% Upper | Wald p-Value | Pct of Total |
|---|---|---|---|---|---|---|---|
| Participant ID | 0.673279 | 0.01348 | 0.000823 | 0.011867 | 0.015094 | <.0001 | 40.237 |
| Residual | | 0.020022 | 0.000522 | 0.019037 | 0.021086 | | 59.763 |
| Total | | 0.033502 | 0.000887 | 0.031829 | 0.035311 | | 100 |

*Abbreviations:* BMI: Body mass index; CN: Cognitively normal; DX: Diagnosis at baseline; EF: Executive Function; EMCI: Early mild cognitive impairment; EntCtx: Entorhinal cortex volume; GlycA: Glycoprotein acetyls; ID: Identification; LMCI: Late mild cognitive impairment; SMC: Significant memory concerns.

**Supplementary Table S2. Baseline GlycA level was associated with EF and MEM in the continuous follow-up years in participants, only in participants diagnosis with LMCI at baseline.**

| Parameters | Year | p-value | | | | | Spearman rho (ρ) | | | | |
|---|---|---|---|---|---|---|---|---|---|---|---|
| | | CN | SMC | EMCI | LMCI | AD | CN | SMC | EMCI | LMCI | AD |
| EF | 1 | 0.301 | 0.994 | 0.512 | 0.153 | 0.884 | 0.057 | -0.002 | 0.041 | -0.069 | 0.01 |
| | 2 | 0.935 | 0.255 | 0.145 | 0.108 | 0.961 | 0.005 | -0.132 | 0.098 | -0.084 | -0.004 |
| | 3 | 0.05 | 0.019 | 0.692 | 0.036 | 0.873 | 0.145 | -0.886 | -0.029 | -0.119 | 0.1 |
| | 4 | 0.519 | 0.442 | 0.044 | 0.001 | NA | 0.047 | 0.123 | 0.164 | -0.236 | -1 |
| | 5 | 0.246 | 0.654 | 0.866 | 0.003 | NA | 0.109 | 0.104 | 0.018 | -0.265 | NA |
| | 6 | 0.587 | 0.467 | 0.884 | 0 | NA | 0.044 | 0.12 | 0.019 | -0.381 | NA |
| | 7 | 0.972 | NA | 0.065 | 0.002 | NA | 0.003 | NA | -0.26 | -0.314 | NA |
| | 8 | 0.286 | NA | 0.866 | 0.001 | NA | -0.116 | NA | -0.031 | -0.391 | NA |
| | 9 | 0.886 | NA | 0.829 | 0.037 | NA | -0.021 | NA | 0.079 | -0.349 | NA |
| | 10 | 0.804 | NA | NA | 0.459 | NA | 0.042 | NA | NA | -0.207 | NA |
| | 11 | 0.109 | NA | NA | 0.747 | NA | 0.284 | NA | NA | -0.2 | NA |
| | 12 | 0.332 | NA | NA | 0.667 | NA | 0.207 | NA | NA | 0.5 | NA |
| | 13 | 0.856 | NA | NA | NA | NA | -0.039 | NA | NA | NA | NA |
| MEM | 1 | 0.695 | 0.69 | 0.381 | 0.236 | 0.242 | -0.022 | -0.084 | -0.055 | -0.057 | -0.079 |
| | 2 | 0.232 | 0.43 | 0.376 | 0.496 | 0.931 | -0.068 | -0.091 | -0.06 | -0.036 | -0.008 |
| | 3 | 0.255 | NA | 0.251 | 0.738 | 0.266 | 0.084 | 1 | -0.083 | -0.019 | -0.543 |
| | 4 | 0.36 | 0.131 | 0.95 | 0.459 | NA | 0.066 | 0.24 | -0.005 | -0.053 | -1 |
| | 5 | 0.147 | 0.713 | 0.158 | 0.036 | NA | 0.136 | -0.081 | -0.146 | -0.183 | NA |
| | 6 | 0.15 | 0.179 | 0.655 | 0.037 | NA | 0.115 | -0.217 | 0.058 | -0.186 | NA |
| | 7 | 0.11 | NA | 0.896 | 0.019 | NA | 0.146 | NA | -0.018 | -0.235 | NA |
| | 8 | 0.244 | NA | 0.343 | 0.036 | NA | 0.126 | NA | 0.171 | -0.262 | NA |
| | 9 | 0.668 | NA | 0.385 | 0.099 | NA | 0.062 | NA | -0.309 | -0.272 | NA |
| | 10 | 0.097 | NA | NA | 0.520 | NA | 0.273 | NA | NA | -0.174 | NA |
| | 11 | 0.134 | NA | NA | 0.266 | NA | 0.266 | NA | NA | -0.543 | NA |
| | 12 | 0.198 | NA | NA | NA | NA | -0.272 | NA | NA | 1 | NA |
| | 13 | NA | NA | NA | NA | NA | NA | NA | NA | NA | NA |

*Abbreviations:* AD: Alzheimer's disease; CN: Cognitively normal; EF: Executive Function; EMCI: Early mild cognitive impairment; GlycA: Glycoprotein acetyls; LMCI: Late mild cognitive impairment; MEM: Memory; SMC: Significant memory concerns.

**Supplementary Table S3. Linear mixed models to evaluate the relationships between GlycA and markers for cognition and structural atrophy.**

| Feature | Model 1 | Model 2 | Model 3 | Model 4 |
|---|---|---|---|---|
| Outcome (Y) | Longitudinal EF | Longitudinal MEM | Longitudinal MEM | Longitudinal EntCtx (ICV and magnet type adjusted) |
| Main Predictor ($X_1$) | Baseline GlycA (medication adjusted) | Baseline GlycA (medication adjusted) | Baseline GlycA (medication adjusted) | Baseline GlycA (medication adjusted) |
| Other covariates ($X_n$) | Baseline EF,<br>Month follow up,<br>Age at screening,<br>BMI at visit,<br>Sex,<br>The presence of ApoE4,<br>Education. | Baseline MEM,<br>Month follow up,<br>Age at screening,<br>BMI at visit,<br>Sex,<br>The presence of ApoE4,<br>Education. | Baseline MEM,<br>Month follow up,<br>Age at screening,<br>BMI at visit,<br>Sex,<br>The presence of ApoE4,<br>Education.<br>DX at baseline. | Baseline EntCtx (ICV and magnet type adjusted),<br>Month follow up,<br>Age at screening,<br>BMI at visit,<br>Sex,<br>The presence of ApoE4,<br>Education. |
| Random effect | Participant ID | Participant ID | Participant ID | Participant ID |
| Stratified by | DX (diagnosis status) at baseline | DX at baseline | | DX at baseline |
| p and $\beta_{estimate}$ for the main predictor | CN: β= 0.01, p=0.932<br>SMC: β= -0.33, p=0.335<br>EMCI: β= 0.08, p=0.661<br>LMCI: β= -0.45, p=0.001<br>AD: β= -0.02, p=0.902 | CN: β= -0.03, p=0.721<br>SMC: β= -0.04, p=0.812<br>EMCI: β= -0.07, p=0.453<br>LMCI: β= -0.14, p=0.063<br>AD: β= -0.04, p=0.611 | β= -0.09, p=0.037 | CN: β= -0.04, p=0.168<br>SMC: β= 0.03, p=0.708<br>EMCI: β= -0.02, p=0.488<br>LMCI: β= -0.10, p<0.001<br>AD: β= 0.02, p=0.485 |

*Abbreviations:* AD: Alzheimer's disease; BMI: Body mass index; CN: Cognitively normal; DX: Diagnosis at baseline; EF: Executive Function; EMCI: Early mild cognitive impairment; EntCtx: Entorhinal cortex volume; GlycA: Glycoprotein acetyls; ICV: Intracranial volume; LMCI: Late mild cognitive impairment; MEM: Memory; SMC: Significant memory concerns.

**Supplementary Table S4. Baseline GlycA level was not associated with other brain regional volumes, other than entorhinal cortex, in the continuous follow up years in participants regardless of participant diagnosis at baseline.**

| Parameters | Year | p-value | | | | | Spearman rho (ρ) | | | | |
|---|---|---|---|---|---|---|---|---|---|---|---|
| | | CN | SMC | EMCI | LMCI | AD | CN | SMC | EMCI | LMCI | AD |
| Bilateral total entorhinal cortex volume | 1 | 0.848 | 0.270 | 0.580 | 0.098 | 0.585 | -0.011 | 0.240 | -0.035 | -0.081 | 0.039 |
| | 2 | 0.281 | 0.635 | 0.443 | 0.011 | 0.671 | -0.064 | 0.060 | -0.054 | -0.140 | -0.041 |
| | 3 | 0.831 | NA | 0.410 | 0.185 | NA | 0.018 | NA | 0.092 | -0.097 | NA |
| | 4 | 0.723 | 0.217 | 0.293 | 0.040 | NA | -0.028 | 0.228 | -0.104 | -0.165 | NA |
| | 5 | 0.425 | 0.638 | 0.182 | 0.115 | NA | 0.090 | -0.109 | -0.265 | -0.172 | NA |
| | 6 | 0.433 | 0.633 | 0.508 | 0.024 | NA | 0.069 | -0.091 | -0.100 | -0.224 | NA |
| | 7 | 0.064 | NA | 0.477 | 0.026 | NA | 0.199 | NA | -0.116 | -0.257 | NA |
| | 8 | 0.228 | NA | 0.296 | 0.025 | NA | 0.148 | NA | 0.194 | -0.345 | NA |
| | 9 | 0.027 | NA | 0.519 | 0.321 | NA | 0.410 | NA | 0.218 | -0.275 | NA |
| | 10 | 0.005 | NA | NA | NA | NA | -0.701 | NA | NA | NA | NA |
| | 11 | 0.322 | NA | NA | NA | NA | -0.227 | NA | NA | NA | NA |
| | 12 | 0.348 | NA | NA | NA | NA | -0.228 | NA | NA | NA | NA |
| | 13 | 0.977 | NA | NA | NA | NA | -0.007 | NA | NA | NA | NA |
| Bilateral total cingulate volume | 1 | 0.479 | 0.154 | 0.230 | 0.957 | 0.282 | -0.039 | -0.307 | -0.077 | 0.003 | 0.076 |
| | 2 | 0.080 | 0.831 | 0.766 | 0.909 | 0.841 | -0.104 | 0.027 | 0.021 | -0.006 | -0.019 |
| | 3 | 0.506 | NA | 0.702 | 0.858 | NA | -0.056 | NA | 0.043 | -0.013 | NA |
| | 4 | 0.857 | 0.695 | 0.946 | 0.138 | NA | -0.014 | 0.073 | 0.007 | -0.119 | NA |
| | 5 | 0.490 | 0.750 | 0.710 | 0.247 | NA | -0.078 | 0.074 | 0.075 | -0.127 | NA |
| | 6 | 0.228 | 0.296 | 0.186 | 0.560 | NA | -0.106 | -0.197 | -0.198 | -0.059 | NA |
| | 7 | 0.855 | NA | 0.939 | 0.626 | NA | -0.020 | NA | -0.013 | -0.057 | NA |
| | 8 | 0.693 | NA | 0.397 | 0.266 | NA | 0.049 | NA | 0.158 | -0.176 | NA |
| | 9 | 0.177 | NA | 0.689 | 0.657 | NA | 0.258 | NA | -0.136 | -0.125 | NA |
| | 10 | 0.615 | NA | NA | NA | NA | 0.147 | NA | NA | NA | NA |
| | 11 | 0.598 | NA | NA | NA | NA | 0.122 | NA | NA | NA | NA |
| | 12 | 0.904 | NA | NA | NA | NA | -0.030 | NA | NA | NA | NA |
| | 13 | 0.823 | NA | NA | NA | NA | 0.057 | NA | NA | NA | NA |
| Total cerebral cortex grey matter volume | 1 | 0.901 | 0.865 | 0.323 | 0.118 | 0.287 | 0.007 | -0.038 | -0.063 | 0.077 | 0.075 |
| | 2 | 0.092 | 0.266 | 0.858 | 0.616 | 0.675 | -0.100 | 0.140 | -0.013 | 0.028 | 0.041 |
| | 3 | 0.486 | NA | 0.904 | 0.371 | NA | 0.059 | NA | -0.014 | -0.066 | NA |

| | | | | | | | | | | | |
|---|---|---|---|---|---|---|---|---|---|---|---|
| | 4 | 0.937 | 0.823 | 0.824 | 0.101 | NA | -0.006 | 0.042 | 0.022 | -0.132 | NA |
| | 5 | 0.207 | 0.699 | 0.585 | 0.113 | NA | 0.143 | -0.090 | -0.110 | -0.173 | NA |
| | 6 | 0.362 | 0.838 | 0.652 | 0.436 | NA | -0.080 | -0.039 | 0.068 | -0.079 | NA |
| | 7 | 0.885 | NA | 0.876 | 0.488 | NA | -0.016 | NA | 0.026 | -0.081 | NA |
| | 8 | 0.245 | NA | 0.591 | 0.033 | NA | 0.143 | NA | 0.100 | -0.331 | NA |
| | 9 | 0.041 | NA | 0.519 | 0.182 | NA | 0.381 | NA | -0.218 | -0.364 | NA |
| | 10 | 0.748 | NA | NA | NA | NA | 0.095 | NA | NA | NA | NA |
| | 11 | 0.239 | NA | NA | NA | NA | 0.269 | NA | NA | NA | NA |
| | 12 | 0.226 | NA | NA | NA | NA | 0.291 | NA | NA | NA | NA |
| | 13 | 0.627 | NA | NA | NA | NA | 0.123 | NA | NA | NA | NA |
| Bilateral total frontal lobe volume | 1 | 0.770 | 0.383 | 0.201 | 0.014 | 0.884 | -0.016 | -0.191 | -0.082 | 0.121 | 0.010 |
| | 2 | 0.181 | 0.166 | 0.912 | 0.078 | 0.756 | -0.079 | 0.174 | 0.008 | 0.098 | 0.030 |
| | 3 | 0.889 | NA | 0.673 | 0.428 | NA | 0.012 | NA | -0.047 | 0.058 | NA |
| | 4 | 0.497 | 0.732 | 0.894 | 0.106 | NA | -0.054 | 0.064 | 0.013 | -0.130 | NA |
| | 5 | 0.797 | 0.548 | 0.873 | 0.239 | NA | 0.029 | 0.139 | -0.032 | -0.129 | NA |
| | 6 | 0.300 | 0.638 | 0.932 | 0.513 | NA | -0.091 | 0.090 | 0.013 | 0.066 | NA |
| | 7 | 0.582 | NA | 0.573 | 0.508 | NA | -0.060 | NA | -0.092 | -0.078 | NA |
| | 8 | 0.854 | NA | 0.406 | 0.185 | NA | 0.023 | NA | 0.155 | -0.209 | NA |
| | 9 | 0.080 | NA | 0.631 | 0.098 | NA | 0.331 | NA | -0.164 | -0.443 | NA |
| | 10 | 0.771 | NA | NA | NA | NA | 0.086 | NA | NA | NA | NA |
| | 11 | 0.263 | NA | NA | NA | NA | 0.256 | NA | NA | NA | NA |
| | 12 | 0.311 | NA | NA | NA | NA | 0.246 | NA | NA | NA | NA |
| | 13 | 0.526 | NA | NA | NA | NA | 0.160 | NA | NA | NA | NA |
| Bilateral mean grey matter volume | 1 | 0.921 | 0.989 | 0.315 | 0.112 | 0.386 | 0.006 | 0.003 | -0.064 | 0.078 | 0.061 |
| | 2 | 0.072 | 0.275 | 0.816 | 0.614 | 0.588 | -0.106 | 0.137 | -0.016 | 0.028 | 0.052 |
| | 3 | 0.529 | NA | 0.925 | 0.399 | NA | 0.053 | NA | -0.011 | -0.062 | NA |
| | 4 | 0.908 | 0.663 | 0.788 | 0.094 | NA | -0.009 | 0.082 | 0.027 | -0.135 | NA |
| | 5 | 0.206 | 0.552 | 0.579 | 0.089 | NA | 0.143 | -0.138 | -0.112 | -0.186 | NA |
| | 6 | 0.327 | 0.986 | 0.816 | 0.428 | NA | -0.086 | 0.003 | 0.035 | -0.080 | NA |
| | 7 | 0.795 | NA | 0.860 | 0.437 | NA | -0.028 | NA | 0.029 | -0.091 | NA |
| | 8 | 0.258 | NA | 0.561 | 0.032 | NA | 0.139 | NA | 0.109 | -0.332 | NA |
| | 9 | 0.043 | NA | 0.519 | 0.173 | NA | 0.378 | NA | -0.218 | -0.371 | NA |
| | 10 | 0.771 | NA | NA | NA | NA | 0.086 | NA | NA | NA | NA |
| | 11 | 0.243 | NA | NA | NA | NA | 0.266 | NA | NA | NA | NA |

| | | | | | | | | | | | |
|---|---|---|---|---|---|---|---|---|---|---|---|
| | 12 | 0.226 | NA | NA | NA | NA | 0.291 | NA | NA | NA | NA |
| | 13 | 0.723 | NA | NA | NA | NA | 0.090 | NA | NA | NA | NA |
| Bilateral mean hippocampus volume | 1 | 0.858 | 0.240 | 0.252 | 0.236 | 0.881 | 0.010 | 0.255 | -0.073 | 0.058 | 0.011 |
| | 2 | 0.147 | 0.266 | 0.952 | 0.221 | 0.391 | -0.086 | -0.140 | 0.004 | -0.068 | -0.083 |
| | 3 | 0.915 | NA | 0.766 | 0.185 | NA | 0.009 | NA | 0.033 | -0.097 | NA |
| | 4 | 0.326 | 0.168 | 0.722 | 0.083 | NA | 0.078 | 0.254 | 0.035 | -0.139 | NA |
| | 5 | 0.253 | 0.827 | 0.885 | 0.189 | NA | 0.129 | 0.051 | 0.029 | -0.144 | NA |
| | 6 | 0.121 | 0.545 | 0.881 | 0.502 | NA | 0.136 | 0.115 | -0.023 | -0.068 | NA |
| | 7 | 0.057 | NA | 0.290 | 0.113 | NA | 0.205 | NA | -0.172 | -0.185 | NA |
| | 8 | 0.041 | NA | 0.693 | 0.036 | NA | 0.249 | NA | 0.074 | -0.325 | NA |
| | 9 | 0.101 | NA | 0.853 | 0.147 | NA | 0.311 | NA | -0.064 | -0.393 | NA |
| | 10 | 0.523 | NA | NA | NA | NA | 0.187 | NA | NA | NA | NA |
| | 11 | 0.162 | NA | NA | NA | NA | 0.317 | NA | NA | NA | NA |
| | 12 | 0.254 | NA | NA | NA | NA | 0.275 | NA | NA | NA | NA |
| | 13 | 0.185 | NA | NA | NA | NA | 0.327 | NA | NA | NA | NA |
| Bilateral total lateral temporal lobe volume | 1 | 0.885 | 0.474 | 0.086 | 0.243 | 0.676 | -0.008 | -0.157 | -0.110 | 0.057 | 0.029 |
| | 2 | 0.031 | 0.234 | 0.259 | 0.431 | 0.170 | -0.127 | 0.150 | -0.079 | -0.044 | 0.132 |
| | 3 | 0.438 | NA | 0.812 | 0.093 | NA | 0.065 | NA | 0.027 | -0.123 | NA |
| | 4 | 0.236 | 0.608 | 0.510 | 0.091 | NA | 0.094 | 0.096 | -0.065 | -0.136 | NA |
| | 5 | 0.027 | 0.253 | 0.072 | 0.113 | NA | 0.248 | -0.261 | -0.352 | -0.173 | NA |
| | 6 | 0.941 | 0.135 | 0.482 | 0.089 | NA | 0.007 | -0.279 | -0.106 | -0.170 | NA |
| | 7 | 0.330 | NA | 0.935 | 0.229 | NA | 0.106 | NA | 0.013 | -0.141 | NA |
| | 8 | 0.081 | NA | 0.955 | 0.084 | NA | 0.213 | NA | 0.011 | -0.269 | NA |
| | 9 | 0.086 | NA | 0.650 | 0.308 | NA | 0.325 | NA | -0.155 | -0.282 | NA |
| | 10 | 0.342 | NA | NA | NA | NA | 0.275 | NA | NA | NA | NA |
| | 11 | 0.987 | NA | NA | NA | NA | 0.004 | NA | NA | NA | NA |
| | 12 | 0.892 | NA | NA | NA | NA | -0.033 | NA | NA | NA | NA |
| | 13 | 0.791 | NA | NA | NA | NA | -0.067 | NA | NA | NA | NA |
| Bilateral total medial temporal lobe volume | 1 | 0.312 | 0.455 | 0.256 | 0.988 | 0.554 | -0.056 | 0.164 | -0.073 | 0.001 | 0.042 |
| | 2 | 0.021 | 0.993 | 0.136 | 0.507 | 0.353 | -0.136 | -0.001 | -0.104 | -0.037 | 0.090 |
| | 3 | 0.541 | NA | 0.389 | 0.187 | NA | 0.052 | NA | 0.097 | -0.096 | NA |
| | 4 | 0.836 | 0.324 | 0.876 | 0.024 | NA | -0.016 | 0.183 | 0.015 | -0.181 | NA |
| | 5 | 0.019 | 0.486 | 0.494 | 0.060 | NA | 0.261 | -0.161 | -0.137 | -0.205 | NA |

| | | | | | | | | | | | |
|---|---|---|---|---|---|---|---|---|---|---|---|
| | 6 | 0.631 | 0.670 | 0.809 | 0.090 | NA | 0.042 | -0.081 | 0.037 | -0.170 | NA |
| | 7 | 0.087 | NA | 0.840 | 0.077 | NA | 0.185 | NA | -0.033 | -0.205 | NA |
| | 8 | 0.046 | NA | 0.404 | 0.067 | NA | 0.242 | NA | 0.155 | -0.286 | NA |
| | 9 | 0.010 | NA | 0.894 | 0.012 | NA | 0.468 | NA | -0.046 | -0.629 | NA |
| | 10 | 0.455 | NA | NA | NA | NA | -0.218 | NA | NA | NA | NA |
| | 11 | 0.268 | NA | NA | NA | NA | 0.253 | NA | NA | NA | NA |
| | 12 | 0.737 | NA | NA | NA | NA | -0.083 | NA | NA | NA | NA |
| | 13 | 0.657 | NA | NA | NA | NA | 0.113 | NA | NA | NA | NA |
| Bilateral total occipital lobe volume | 1 | 0.917 | 0.500 | 0.092 | 0.619 | 0.459 | 0.006 | 0.148 | -0.108 | 0.024 | 0.052 |
| | 2 | 0.186 | 0.245 | 0.527 | 0.796 | 0.266 | -0.078 | 0.146 | 0.044 | 0.014 | 0.108 |
| | 3 | 0.690 | NA | 0.634 | 0.792 | NA | 0.034 | NA | 0.053 | -0.019 | NA |
| | 4 | 0.455 | 0.748 | 0.789 | 0.028 | NA | -0.059 | 0.060 | 0.026 | -0.177 | NA |
| | 5 | 0.645 | 0.430 | 0.596 | 0.282 | NA | 0.052 | -0.182 | -0.107 | -0.118 | NA |
| | 6 | 0.073 | 0.682 | 0.402 | 0.779 | NA | -0.157 | 0.078 | 0.127 | 0.028 | NA |
| | 7 | 0.367 | NA | 0.470 | 0.587 | NA | -0.098 | NA | 0.118 | -0.064 | NA |
| | 8 | 0.633 | NA | 0.398 | 0.151 | NA | 0.059 | NA | 0.157 | -0.226 | NA |
| | 9 | 0.336 | NA | 0.417 | 0.173 | NA | 0.185 | NA | -0.273 | -0.371 | NA |
| | 10 | 0.829 | NA | NA | NA | NA | 0.064 | NA | NA | NA | NA |
| | 11 | 0.205 | NA | NA | NA | NA | 0.288 | NA | NA | NA | NA |
| | 12 | 0.557 | NA | NA | NA | NA | -0.144 | NA | NA | NA | NA |
| | 13 | 0.868 | NA | NA | NA | NA | -0.042 | NA | NA | NA | NA |
| Bilateral total parietal lobe volume | 1 | 0.872 | 0.819 | 0.319 | 0.140 | 0.248 | 0.009 | 0.050 | -0.064 | 0.072 | 0.081 |
| | 2 | 0.055 | 0.335 | 0.742 | 0.684 | 0.944 | -0.113 | 0.121 | -0.023 | 0.023 | 0.007 |
| | 3 | 0.940 | NA | 0.159 | 0.161 | NA | 0.006 | NA | -0.157 | -0.103 | NA |
| | 4 | 0.687 | 0.884 | 0.403 | 0.133 | NA | -0.032 | 0.027 | 0.082 | -0.121 | NA |
| | 5 | 0.396 | 0.737 | 0.652 | 0.058 | NA | 0.096 | -0.078 | 0.091 | -0.206 | NA |
| | 6 | 0.116 | 0.864 | 0.234 | 0.362 | NA | -0.138 | 0.033 | 0.179 | -0.092 | NA |
| | 7 | 0.185 | NA | 0.782 | 0.601 | NA | -0.143 | NA | 0.045 | -0.061 | NA |
| | 8 | 0.435 | NA | 0.949 | 0.013 | NA | 0.096 | NA | -0.012 | -0.381 | NA |
| | 9 | 0.089 | NA | 0.689 | 0.383 | NA | 0.321 | NA | -0.136 | -0.243 | NA |
| | 10 | 0.946 | NA | NA | NA | NA | -0.020 | NA | NA | NA | NA |
| | 11 | 0.260 | NA | NA | NA | NA | 0.257 | NA | NA | NA | NA |
| | 12 | 0.329 | NA | NA | NA | NA | 0.237 | NA | NA | NA | NA |
| | 13 | 0.311 | NA | NA | NA | NA | 0.253 | NA | NA | NA | NA |

| | | | | | | | | | | | |
|---|---|---|---|---|---|---|---|---|---|---|---|
| Bilateral total sensory motor volume | 1 | 0.801 | 0.556 | 0.304 | 0.104 | 0.413 | -0.014 | -0.129 | -0.066 | 0.080 | 0.058 |
| | 2 | 0.276 | 0.954 | 0.821 | 0.127 | 0.642 | -0.065 | 0.007 | -0.016 | 0.085 | 0.045 |
| | 3 | 0.852 | NA | 0.396 | 0.578 | NA | 0.016 | NA | -0.095 | 0.041 | NA |
| | 4 | 0.409 | 0.998 | 0.181 | 0.917 | NA | -0.065 | 0.000 | 0.132 | -0.008 | NA |
| | 5 | 0.765 | 0.862 | 0.392 | 0.125 | NA | 0.034 | -0.040 | -0.172 | -0.168 | NA |
| | 6 | 0.067 | 0.795 | 0.369 | 0.892 | NA | -0.160 | -0.050 | 0.136 | 0.014 | NA |
| | 7 | 0.106 | NA | 0.252 | 0.637 | NA | -0.174 | NA | 0.186 | -0.055 | NA |
| | 8 | 0.548 | NA | 0.180 | 0.025 | NA | 0.074 | NA | 0.247 | -0.345 | NA |
| | 9 | 0.124 | NA | 0.285 | 0.277 | NA | 0.292 | NA | -0.355 | -0.300 | NA |
| | 10 | 0.958 | NA | NA | NA | NA | -0.015 | NA | NA | NA | NA |
| | 11 | 0.203 | NA | NA | NA | NA | 0.290 | NA | NA | NA | NA |
| | 12 | 0.446 | NA | NA | NA | NA | 0.186 | NA | NA | NA | NA |
| | 13 | 0.414 | NA | NA | NA | NA | 0.205 | NA | NA | NA | NA |
| Bilateral total temporal lobe volume | 1 | 0.516 | 0.747 | 0.142 | 0.376 | 0.662 | -0.036 | -0.071 | -0.094 | 0.044 | 0.031 |
| | 2 | 0.013 | 0.336 | 0.141 | 0.374 | 0.244 | -0.146 | 0.121 | -0.103 | -0.049 | 0.113 |
| | 3 | 0.389 | NA | 0.624 | 0.120 | NA | 0.073 | NA | 0.055 | -0.114 | NA |
| | 4 | 0.479 | 0.375 | 0.657 | 0.051 | NA | 0.056 | 0.165 | -0.044 | -0.157 | NA |
| | 5 | 0.023 | 0.297 | 0.134 | 0.102 | NA | 0.255 | -0.239 | -0.296 | -0.178 | NA |
| | 6 | 0.784 | 0.307 | 0.671 | 0.069 | NA | 0.024 | -0.193 | -0.064 | -0.182 | NA |
| | 7 | 0.205 | NA | 0.830 | 0.167 | NA | 0.137 | NA | -0.035 | -0.161 | NA |
| | 8 | 0.057 | NA | 0.696 | 0.062 | NA | 0.232 | NA | 0.073 | -0.290 | NA |
| | 9 | 0.019 | NA | 0.650 | 0.111 | NA | 0.433 | NA | -0.155 | -0.429 | NA |
| | 10 | 0.615 | NA | NA | NA | NA | 0.147 | NA | NA | NA | NA |
| | 11 | 0.666 | NA | NA | NA | NA | 0.100 | NA | NA | NA | NA |
| | 12 | 0.904 | NA | NA | NA | NA | -0.030 | NA | NA | NA | NA |
| | 13 | 0.735 | NA | NA | NA | NA | 0.086 | NA | NA | NA | NA |
| White Matter Hyperintensities | 1 | 0.505 | 0.248 | 0.929 | 0.050 | 0.212 | -0.037 | -0.251 | -0.006 | 0.097 | 0.088 |
| | 2 | 0.845 | 0.313 | 0.606 | 0.203 | 0.593 | -0.012 | -0.128 | -0.036 | 0.072 | 0.053 |
| | 3 | 0.078 | NA | 0.364 | 0.858 | NA | -0.160 | NA | 0.101 | 0.015 | NA |
| | 4 | 0.484 | 0.467 | 0.746 | 0.560 | NA | 0.070 | 0.136 | 0.032 | 0.073 | NA |
| | 5 | 0.647 | 0.291 | 0.023 | 0.693 | NA | -0.112 | -0.242 | 0.437 | 0.100 | NA |
| | 6 | 0.299 | 0.901 | 0.720 | 0.681 | NA | -0.131 | 0.024 | 0.054 | 0.070 | NA |
| | 7 | 0.019 | NA | 0.960 | 0.747 | NA | -0.361 | NA | -0.008 | 0.052 | NA |

| | | | | | | | | | | |
|---|---|---|---|---|---|---|---|---|---|---|
| 8 | 0.122 | NA | 0.980 | 0.155 | NA | -0.237 | NA | -0.005 | 0.293 | NA |
| 9 | 0.762 | NA | 0.043 | 0.383 | NA | -0.098 | NA | 0.649 | 0.393 | NA |
| 10 | 0.170 | NA | NA | NA | NA | -0.500 | NA | NA | NA | NA |
| 11 | 0.504 | NA | NA | NA | NA | -0.155 | NA | NA | NA | NA |
| 12 | 0.681 | NA | NA | NA | NA | -0.104 | NA | NA | NA | NA |
| 13 | 0.997 | NA | NA | NA | NA | 0.001 | NA | NA | NA | NA |

*Abbreviations:* AD: Alzheimer's disease; CN: Cognitively normal; EMCI: Early mild cognitive impairment; EntCtx: Entorhinal cortex volume; LMCI: Late mild cognitive impairment; SMC: Significant memory concerns.

**Supplementary Table S5. Plasma GlycA was correlated with regional brain volumes in females with AD and LMCI.** Spearman's rank order correlation was performed between the residual of GlycA (log2-transformed, adjusted medications, APOE4, age, BMI, and education), and residuals of whole brain regional volumes (log2-transformed, adjusted intracranial volume (log2 transformed), magnet strength/scan type, APOE4, age, BMI, education); the analysis was performed stratified by sex and diagnosis groups. In addition, linear regressions were performed to evaluate the GlycA-sex interaction stratified by diagnosis status, and GlycA-diagnosis status interaction stratified by sex; both analyses were again done while also adjusting APOE4, age, BMI, education, magnet strength/scan type, and intracranial volume (log2 transformed). FDR correlations were performed for each stratified analysis. Results annotated with * and underlined were significant ($p$<0.05) and passed FDR correction of q=0.2; other highlighted results were significant ($p$<0.05) but did not passed FDR.

| Measurement | Spearman rho (ρ) | | | | | | | | | | *p* (interaction) | | | | | | |
|---|---|---|---|---|---|---|---|---|---|---|---|---|---|---|---|---|---|
| | Male | | | | | Female | | | | | GlycA-sex interaction | | | | | GlycaA-diagnosis interaction | |
| | CN | SMC | EMCI | LMCI | AD | CN | SMC | EMCI | LMCI | AD | CN | SMC | EMCI | LMCI | AD | Male | Female |
| Bilateral total frontal lobe volume | 0.025 | -0.061 | 0.017 | 0.032 | 0.179 | 0.056 | **-0.337 *** | -0.041 | 0.028 | **-0.343 *** | 0.457 | 0.312 | 0.734 | 0.532 | **<0.001 *** | 0.569 | 0.021 |
| Total cerebral cortex grey matter volume | 0.028 | -0.056 | -0.066 | 0.017 | 0.096 | 0.118 | **-0.329 *** | -0.023 | -0.002 | **-0.269 *** | 0.379 | 0.424 | 0.389 | 0.397 | **0.008 *** | 0.357 | 0.022 |
| Bilateral mean grey matter volume | 0.024 | -0.044 | -0.057 | 0.018 | 0.101 | 0.114 | **-0.341 *** | -0.024 | -0.005 | **-0.267 *** | 0.352 | 0.428 | 0.443 | 0.341 | **0.007 *** | 0.374 | 0.031 |
| Bilateral total parietal lobe volume | -0.002 | -0.010 | -0.133 | 0.030 | 0.040 | 0.071 | -0.254 | 0.054 | 0.022 | **-0.222 *** | 0.355 | 0.617 | 0.068 | 0.790 | 0.094 | 0.182 | 0.207 |
| Bilateral total sensory motor volume | -0.046 | -0.028 | 0.065 | -0.006 | 0.080 | 0.036 | -0.246 | 0.012 | 0.116 | **-0.214 *** | 0.084 | 0.705 | 0.796 | 0.181 | **0.020 *** | 0.493 | 0.105 |
| Bilateral mean hippocampus volume | 0.039 | -0.049 | -0.075 | -0.094 | -0.183 | 0.081 | 0.151 | -0.156 | **-0.186 *** | 0.001 | 0.662 | 0.276 | 0.790 | 0.183 | 0.082 | 0.123 | 0.111 |
| Bilateral total entorhinal cortex volume | 0.042 | -0.275 | 0.015 | 0.019 | -0.123 | 0.108 | 0.049 | -0.111 | **-0.163 *** | 0.104 | 0.631 | 0.130 | 0.544 | 0.019 | 0.112 | 0.118 | 0.040 |
| Bilateral total cingulate volume | 0.077 | -0.099 | -0.069 | 0.001 | 0.039 | 0.086 | -0.187 | -0.079 | 0.002 | -0.153 | 0.748 | 0.469 | 0.320 | 0.993 | 0.516 | 0.383 | 0.088 |
| Bilateral total lateral temporal lobe volume | 0.018 | -0.103 | -0.080 | 0.048 | 0.085 | 0.165 | -0.185 | -0.045 | -0.075 | -0.103 | 0.268 | 0.526 | 0.474 | 0.027 | 0.200 | 0.260 | 0.041 |
| Bilateral total temporal lobe volume | 0.038 | -0.172 | -0.065 | 0.011 | 0.057 | 0.169 | -0.173 | -0.044 | -0.087 | -0.077 | 0.318 | 0.863 | 0.438 | 0.022 | 0.294 | 0.170 | 0.027 |
| Bilateral total occipital lobe volume | 0.087 | 0.163 | -0.069 | -0.020 | -0.037 | 0.063 | -0.101 | 0.006 | 0.102 | -0.076 | 0.405 | 0.290 | 0.368 | 0.975 | 0.580 | 0.336 | 0.793 |
| Bilateral total medial temporal lobe volume | 0.094 | -0.275 | -0.064 | -0.046 | -0.025 | 0.106 | -0.053 | -0.044 | -0.09 | -0.032 | 0.811 | 0.324 | 0.533 | 0.065 | 0.740 | 0.154 | 0.122 |

*Abbreviations:* AD: Alzheimer's disease; BMI: Body mass index; CN: Cognitively normal; EMCI: Early mild cognitive impairment; EntCtx: Entorhinal cortex volume; FDR: False discovery rate: GlycA: Glycoprotein acetyls; LMCI: Late mild cognitive impairment; SMC: Significant memory concerns.

**Supplementary Table S6. The sex- and diagnosis groups-specific correlation between plasma GlycA and regional brain volumes (volumes and thicknesses in the left/right hemispheres and the whole brain thicknesses).** Spearman's rank order correlation was performed between the residual of GlycA (log2-transformed, adjusted medications, APOE4, age, BMI, and education, stratified by sex and diagnosis groups) and residuals of brain regional volumes (log2-transformed, adjusted intracranial volume (log2-transformed), magnet strength/scan type (Mag), APOE4, age, BMI, education, stratified by sex and diagnosis groups); the analysis was performed stratified by sex and diagnosis groups. In addition, linear regressions were performed to evaluate the GlycA-sex interaction stratified by diagnosis status, and GlycA-diagnosis status interaction stratified by sex. Results annotated underlined were significant ($p$<0.05) and passed FDR correction of q=0.2; other highlighted results were significant ($p$<0.05) but did not passed FDR.

| Measurement | | | Spearman rho (ρ) | | | | | | | | | | *p* (interaction) | | | | | | |
|---|---|---|---|---|---|---|---|---|---|---|---|---|---|---|---|---|---|---|---|
| | | | Male | | | | | Female | | | | | GlycA-sex interaction | | | | | GlycA-Diagnosis interaction | |
| | | | CN | SMC | EMCI | LMCI | AD | CN | SMC | EMCI | LMCI | AD | CN | SMC | EMCI | LMCI | AD | Male | Female |
| Right total frontal lobe volume | Right | Volume | 0.039 | -0.070 | 0.045 | 0.032 | 0.228 | 0.088 | -0.425 | -0.056 | 0.022 | -0.299 | 0.445 | 0.263 | 0.511 | 0.650 | 0.000 | 0.371 | 0.008 |
| Right cerebral cortex grey matter volume | Right | Volume | 0.024 | -0.079 | -0.043 | 0.014 | 0.110 | 0.119 | -0.338 | -0.023 | 0.004 | -0.282 | 0.314 | 0.528 | 0.588 | 0.626 | 0.004 | 0.457 | 0.007 |
| Right mean grey matter volume | Right | Volume | 0.018 | -0.060 | -0.031 | 0.014 | 0.112 | 0.122 | -0.328 | -0.024 | 0.003 | -0.277 | 0.286 | 0.506 | 0.674 | 0.590 | 0.003 | 0.469 | 0.011 |
| Right total parietal lobe volume | Right | Volume | -0.020 | -0.048 | -0.091 | 0.053 | 0.022 | 0.095 | -0.206 | 0.038 | 0.009 | -0.234 | 0.207 | 0.962 | 0.113 | 0.760 | 0.073 | 0.178 | 0.058 |
| Right total sensory motor volume | Right | Volume | -0.070 | -0.013 | -0.002 | -0.013 | 0.085 | 0.031 | -0.227 | -0.008 | 0.104 | -0.229 | 0.074 | 0.649 | 0.815 | 0.148 | 0.009 | 0.415 | 0.067 |
| Right total cingulate volume | Right | Volume | 0.093 | 0.063 | -0.127 | -0.019 | 0.032 | 0.063 | 0.063 | -0.025 | -0.024 | -0.151 | 0.486 | 0.867 | 0.187 | 0.872 | 0.544 | 0.296 | 0.341 |
| Right total lateral temporal lobe volume | Right | Volume | 0.000 | -0.017 | 0.013 | 0.027 | 0.085 | 0.146 | -0.105 | -0.046 | -0.072 | -0.096 | 0.224 | 0.543 | 0.921 | 0.274 | 0.156 | 0.841 | 0.099 |
| Right total temporal lobe volume | Right | Volume | 0.022 | -0.124 | -0.024 | -0.003 | 0.049 | 0.163 | -0.162 | -0.030 | -0.078 | -0.095 | 0.236 | 0.743 | 0.814 | 0.209 | 0.256 | 0.608 | 0.051 |
| Right total medial temporal lobe volume | Right | Volume | 0.076 | -0.246 | -0.063 | -0.043 | -0.045 | 0.121 | -0.130 | 0.006 | -0.066 | -0.065 | 0.549 | 0.612 | 0.426 | 0.208 | 0.752 | 0.229 | 0.104 |
| Right total occipital lobe volume | Right | Volume | 0.085 | 0.092 | -0.079 | -0.040 | -0.051 | 0.057 | -0.059 | -0.002 | 0.114 | -0.043 | 0.301 | 0.543 | 0.410 | 0.568 | 0.469 | 0.273 | 0.758 |
| Right hippocampus volume | Right | Volume | 0.031 | 0.039 | -0.056 | -0.081 | -0.114 | 0.076 | 0.199 | -0.189 | -0.181 | 0.021 | 0.507 | 0.394 | 0.535 | 0.159 | 0.232 | 0.327 | 0.073 |
| Right entorhinal cortex volume | Right | Volume | 0.010 | -0.303 | -0.015 | -0.003 | -0.044 | 0.118 | -0.045 | -0.110 | -0.184 | 0.052 | 0.253 | 0.368 | 0.641 | 0.018 | 0.272 | 0.240 | 0.037 |
| Left total frontal lobe volume | Left | Volume | 0.001 | -0.029 | -0.030 | 0.032 | 0.126 | 0.032 | -0.210 | -0.044 | 0.021 | -0.357 | 0.499 | 0.429 | 0.939 | 0.446 | 0.003 | 0.748 | 0.091 |
| Left cerebral cortex grey matter volume | Left | Volume | 0.037 | -0.056 | -0.095 | 0.021 | 0.081 | 0.110 | -0.310 | -0.020 | -0.015 | -0.235 | 0.463 | 0.339 | 0.245 | 0.238 | 0.036 | 0.292 | 0.074 |
| Left mean grey matter volume | Left | Volume | 0.031 | -0.030 | -0.086 | 0.022 | 0.084 | 0.109 | -0.278 | -0.008 | -0.017 | -0.234 | 0.443 | 0.363 | 0.272 | 0.183 | 0.036 | 0.307 | 0.100 |
| Left total parietal lobe volume | Left | Volume | 0.004 | 0.048 | -0.172 | 0.003 | 0.063 | 0.040 | -0.336 | 0.053 | 0.026 | -0.195 | 0.617 | 0.294 | 0.055 | 0.851 | 0.188 | 0.217 | 0.394 |
| Left total sensory motor volume | Left | Volume | -0.032 | -0.052 | 0.123 | 0.001 | 0.025 | 0.042 | -0.179 | 0.036 | 0.120 | -0.137 | 0.137 | 0.809 | 0.801 | 0.271 | 0.073 | 0.655 | 0.255 |
| Left total lateral temporal lobe volume | Left | Volume | 0.045 | -0.115 | -0.120 | 0.050 | 0.098 | 0.171 | -0.225 | -0.025 | -0.068 | -0.105 | 0.367 | 0.548 | 0.159 | 0.003 | 0.434 | 0.061 | 0.017 |
| Left total temporal lobe volume | Left | Volume | 0.060 | -0.233 | -0.103 | 0.011 | 0.059 | 0.165 | -0.160 | -0.050 | -0.090 | -0.074 | 0.465 | 0.981 | 0.220 | 0.002 | 0.498 | 0.057 | 0.018 |
| Left total occipital lobe volume | Left | Volume | 0.087 | 0.164 | -0.028 | -0.003 | -0.005 | 0.060 | -0.102 | -0.008 | 0.061 | -0.063 | 0.609 | 0.161 | 0.374 | 0.569 | 0.766 | 0.456 | 0.841 |
| Left total cingulate volume | Left | Volume | 0.012 | -0.079 | -0.013 | 0.030 | 0.024 | 0.091 | -0.394 | -0.089 | 0.031 | -0.042 | 0.882 | 0.303 | 0.704 | 0.823 | 0.758 | 0.744 | 0.018 |
| Left hippocampus volume | Left | Volume | 0.050 | -0.200 | -0.072 | -0.098 | -0.222 | 0.064 | 0.102 | -0.107 | -0.176 | -0.004 | 0.893 | 0.202 | 0.915 | 0.259 | 0.048 | 0.067 | 0.255 |
| Left total medial temporal lobe volume | Left | Volume | 0.088 | -0.218 | -0.050 | -0.062 | -0.003 | 0.077 | 0.033 | -0.089 | -0.101 | 0.003 | 0.845 | 0.213 | 0.716 | 0.036 | 0.754 | 0.294 | 0.217 |
| Left entorhinal cortex volume | Left | Volume | 0.085 | -0.247 | 0.029 | 0.016 | -0.194 | 0.033 | 0.165 | -0.083 | -0.099 | 0.151 | 0.750 | 0.063 | 0.517 | 0.063 | 0.091 | 0.117 | 0.194 |
| bilateral mean frontal lobe thickness | Whole | Thickness | -0.043 | 0.160 | -0.045 | 0.018 | 0.188 | -0.148 | -0.102 | 0.022 | 0.032 | -0.224 | 0.437 | 0.515 | 0.867 | 0.690 | 0.002 | 0.426 | 0.147 |
| bilateral mean parietal lobe thickness | Whole | Thickness | 0.014 | 0.187 | -0.143 | 0.080 | 0.159 | -0.005 | 0.072 | 0.116 | 0.051 | -0.140 | 0.995 | 0.463 | 0.037 | 0.792 | 0.038 | 0.157 | 0.668 |
| bilateral mean grey matter thickness | Whole | Thickness | -0.026 | 0.061 | -0.134 | 0.047 | 0.155 | -0.075 | -0.072 | 0.073 | 0.014 | -0.137 | 0.731 | 0.682 | 0.130 | 0.793 | 0.017 | 0.231 | 0.504 |
| bilateral mean occipital lobe thickness | Whole | Thickness | 0.096 | 0.256 | -0.136 | 0.007 | 0.134 | 0.036 | -0.266 | 0.169 | 0.105 | -0.103 | 0.573 | 0.013 | 0.007 | 0.166 | 0.064 | 0.043 | 0.061 |
| bilateral mean lateral temporal lobe thickness | Whole | Thickness | -0.068 | -0.025 | -0.199 | 0.063 | 0.130 | -0.054 | 0.001 | 0.100 | -0.074 | -0.085 | 0.896 | 0.782 | 0.041 | 0.069 | 0.143 | 0.039 | 0.786 |
| bilateral mean sensory motor thickness | Whole | Thickness | -0.034 | -0.021 | -0.046 | 0.045 | 0.141 | 0.006 | -0.106 | 0.018 | 0.148 | -0.064 | 0.374 | 0.703 | 0.573 | 0.329 | 0.037 | 0.301 | 0.371 |
| bilateral mean temporal lobe thickness | Whole | Thickness | -0.058 | -0.062 | -0.181 | 0.032 | 0.084 | -0.059 | 0.007 | 0.088 | -0.076 | -0.058 | 0.989 | 0.525 | 0.068 | 0.084 | 0.275 | 0.086 | 0.783 |
| bilateral mean medial temporal lobe thickness | Whole | Thickness | -0.024 | -0.220 | -0.135 | -0.014 | 0.012 | -0.059 | -0.012 | 0.055 | -0.080 | -0.036 | 0.832 | 0.357 | 0.251 | 0.190 | 0.598 | 0.363 | 0.868 |
| bilateral mean cingulate thickness | Whole | Thickness | -0.084 | 0.051 | 0.129 | -0.017 | -0.025 | -0.089 | -0.208 | -0.027 | 0.033 | -0.012 | 0.827 | 0.717 | 0.099 | 0.251 | 0.528 | 0.473 | 0.617 |
| bilateral mean entorhinal cortex thickness | Whole | Thickness | 0.027 | -0.416 | -0.056 | 0.037 | -0.052 | -0.070 | 0.007 | -0.038 | -0.141 | 0.050 | 0.312 | 0.113 | 0.943 | 0.014 | 0.332 | 0.164 | 0.451 |
| Right mean frontal lobe thickness | Right | Thickness | -0.014 | 0.165 | 0.005 | -0.010 | 0.247 | -0.146 | -0.041 | -0.021 | 0.031 | -0.244 | 0.490 | 0.615 | 0.404 | 0.608 | 0.000 | 0.147 | 0.128 |
| Right mean cingulate thickness | Right | Thickness | -0.022 | 0.021 | 0.082 | -0.112 | -0.038 | -0.071 | 0.125 | -0.034 | 0.062 | -0.226 | 0.517 | 0.533 | 0.171 | 0.023 | 0.034 | 0.304 | 0.000 |
| Right mean grey matter thickness | Right | Thickness | -0.018 | 0.090 | -0.097 | 0.033 | 0.172 | -0.075 | -0.019 | 0.066 | 0.014 | -0.157 | 0.693 | 0.754 | 0.230 | 0.992 | 0.008 | 0.315 | 0.284 |
| Right mean parietal lobe thickness | Right | Thickness | 0.005 | 0.118 | -0.122 | 0.071 | 0.130 | 0.000 | 0.071 | 0.068 | 0.035 | -0.144 | 0.708 | 0.592 | 0.042 | 0.789 | 0.033 | 0.222 | 0.423 |
| Right mean occipital lobe thickness | Right | Thickness | 0.114 | 0.275 | -0.111 | 0.030 | 0.100 | 0.033 | -0.240 | 0.162 | 0.131 | -0.114 | 0.378 | 0.020 | 0.013 | 0.112 | 0.046 | 0.052 | 0.049 |
| Right mean sensory motor thickness | Right | Thickness | -0.048 | -0.046 | -0.033 | 0.047 | 0.152 | -0.002 | -0.148 | 0.018 | 0.119 | -0.104 | 0.457 | 0.834 | 0.636 | 0.443 | 0.016 | 0.361 | 0.319 |
| Right mean lateral temporal lobe thickness | Right | Thickness | -0.029 | 0.061 | -0.124 | 0.073 | 0.120 | -0.044 | 0.075 | 0.082 | -0.070 | -0.061 | 0.991 | 0.711 | 0.276 | 0.235 | 0.174 | 0.177 | 0.929 |
| Right mean temporal lobe thickness | Right | Thickness | -0.042 | -0.069 | -0.106 | 0.050 | 0.094 | -0.054 | 0.070 | 0.101 | -0.089 | -0.051 | 0.971 | 0.408 | 0.301 | 0.263 | 0.225 | 0.240 | 0.874 |
| Right mean medial temporal lobe thickness | Right | Thickness | -0.050 | -0.225 | -0.101 | -0.004 | 0.064 | -0.047 | 0.102 | 0.086 | -0.101 | -0.049 | 0.918 | 0.216 | 0.541 | 0.445 | 0.299 | 0.504 | 0.776 |
| Right entorhinal cortex thickness | Right | Thickness | 0.027 | -0.420 | -0.014 | 0.020 | -0.001 | -0.069 | 0.084 | 0.024 | -0.107 | -0.009 | 0.329 | 0.070 | 0.956 | 0.095 | 0.996 | 0.327 | 0.955 |
| Left mean frontal lobe thickness | Left | Thickness | -0.056 | 0.100 | -0.088 | 0.046 | 0.114 | -0.146 | -0.203 | 0.045 | 0.041 | -0.189 | 0.431 | 0.445 | 0.614 | 0.776 | 0.023 | 0.656 | 0.210 |
| Left mean parietal lobe thickness | Left | Thickness | 0.024 | 0.172 | -0.140 | 0.078 | 0.165 | -0.020 | 0.045 | 0.127 | 0.073 | -0.121 | 0.707 | 0.375 | 0.045 | 0.795 | 0.074 | 0.146 | 0.882 |

| | | | | | | | | | | | | | | | | | | | |
|---|---|---|---|---|---|---|---|---|---|---|---|---|---|---|---|---|---|---|---|
| Left mean lateral temporal lobe thickness | Left | Thickness | -0.098 | -0.102 | **-0.245** | 0.042 | 0.107 | -0.041 | -0.037 | 0.107 | -0.050 | -0.097 | 0.817 | 0.879 | **0.009** | **0.016** | 0.254 | 0.021 | 0.549 |
| Left mean grey matter thickness | Left | Thickness | -0.029 | -0.002 | -0.158 | 0.049 | 0.122 | -0.068 | -0.088 | 0.072 | 0.009 | -0.095 | 0.773 | 0.629 | 0.080 | 0.570 | 0.063 | 0.176 | 0.779 |
| Left mean occipital lobe thickness | Left | Thickness | 0.077 | 0.213 | -0.140 | -0.030 | 0.145 | 0.033 | -0.251 | 0.162 | 0.067 | -0.072 | 0.897 | **0.017** | **0.007** | 0.366 | 0.165 | 0.080 | 0.170 |
| Left mean temporal lobe thickness | Left | Thickness | -0.072 | -0.142 | **-0.218** | 0.008 | 0.064 | -0.050 | -0.012 | 0.080 | -0.061 | -0.056 | 0.932 | 0.686 | **0.019** | **0.022** | 0.479 | 0.058 | 0.548 |
| Left mean cingulate thickness | Left | Thickness | -0.062 | 0.179 | 0.082 | 0.006 | -0.062 | -0.100 | 0.075 | -0.014 | -0.021 | -0.052 | 0.853 | 0.730 | 0.298 | 0.924 | 0.450 | 0.743 | 0.169 |
| Left mean medial temporal lobe thickness | Left | Thickness | -0.001 | -0.138 | **-0.161** | -0.035 | -0.022 | -0.045 | -0.109 | -0.002 | -0.064 | -0.002 | 0.588 | 0.670 | 0.138 | 0.088 | 0.994 | 0.354 | 0.693 |
| Left mean sensory motor thickness | Left | Thickness | -0.019 | 0.008 | -0.060 | 0.038 | 0.127 | 0.016 | -0.063 | 0.010 | 0.165 | 0.000 | 0.333 | 0.622 | 0.552 | 0.253 | 0.098 | 0.291 | 0.441 |
| Left entorhinal cortex thickness | Left | Thickness | 0.038 | -0.303 | -0.056 | 0.031 | -0.135 | -0.064 | -0.020 | -0.121 | -0.145 | 0.139 | 0.469 | 0.326 | 0.976 | **0.004** | 0.079 | 0.119 | 0.068 |

*Abbreviations:* AD: Alzheimer's disease; BMI: Body mass index; CN: Cognitively normal; EMCI: Early mild cognitive impairment; FDR: False discovery rate: GlycA: Glycoprotein acetyls; LMCI: Late mild cognitive impairment; SMC: Significant memory concerns.

**Supplementary Table S7. The MRI regional brain volumes in all participants.** The original numbers of mean ± standard deviation were reported for each brain measurements. Comparing different diagnosis status within the same measurements, the level sharing the same letters from A-D indicate no significant differences. The significance difference level among different diagnosis status was determined using the Tukey HSD test, using data that were log2-transformed and adjusted for APOE4, sex, age, BMI, education, magnet strength/scan type, and intracranial volume (log2 transformed).

| Brain Volumes | CN (n=361) | | SMC (n=95) | | EMCI (n=279) | | LMCI (n=479) | | AD (n=285) | |
|---|---|---|---|---|---|---|---|---|---|---|
| Bilateral total entorhinal cortex volume | 4056.2 ± 689.3 | A | 4142.5 ± 679.6 | AB | 4042.3 ± 809.6 | B | 3667.9 ± 875.3 | C | 3149 ± 815 | D |
| Bilateral mean hippocampus volume | 3648.7 ± 428.8 | A | 3827.9 ± 480.3 | A | 3684.1 ± 455.8 | B | 3266.7 ± 502.1 | C | 3020.6 ± 485 | D |
| Bilateral total lateral temporal lobe volume | 66659.3 ± 7460.3 | A | 69340 ± 7428 | AB | 68560.6 ± 7975.8 | B | 63181.8 ± 7939.4 | C | 58799.2 ± 8793.4 | D |
| Bilateral total medial temporal lobe volume | 30015.5 ± 3061.1 | A | 30834.6 ± 2987.1 | AB | 30613 ± 3649.9 | B | 28408.9 ± 3593.9 | C | 26215.8 ± 3810.1 | D |
| Bilateral total temporal lobe volume | 96674.8 ± 9822.2 | A | 100174.6 ± 9559.8 | AB | 99173.6 ± 10920.9 | B | 91590.7 ± 10937.4 | C | 85015 ± 12025.5 | D |
| Bilateral total cingulate volume | 17297.9 ± 1984.9 | A | 18348.8 ± 2683.8 | A | 18086.5 ± 2360.5 | AB | 17047 ± 2223.2 | B | 16681 ± 2481.5 | C |
| Total cerebral cortex grey matter volume | 411173.2 ± 39284.3 | A | 428775.2 ± 36155.2 | A | 430354.9 ± 43743.3 | A | 399974.9 ± 42677.5 | B | 385020.9 ± 45453.2 | C |
| Bilateral total frontal lobe volume | 126440.3 ± 12621.1 | A | 130771.9 ± 12795.5 | AB | 131424 ± 13783.6 | A | 123914.3 ± 13774.5 | B | 121325.8 ± 14810.8 | C |
| Bilateral mean grey matter volume | 394647 ± 37904.3 | A | 411165.9 ± 34739.1 | A | 413070.2 ± 41868.8 | A | 383985.8 ± 40965.9 | B | 369522.4 ± 43814.9 | C |
| Bilateral total parietal lobe volume | 78914.4 ± 8676.4 | A | 82533.3 ± 7753.5 | AB | 83412.8 ± 9868.5 | A | 76553.2 ± 9798.1 | B | 72937.7 ± 10477.4 | C |
| Bilateral total sensory motor volume | 45554.7 ± 5211.3 | A | 48050.3 ± 4807.9 | AB | 48822.5 ± 5858.2 | A | 44939.1 ± 5592.7 | AB | 44649.1 ± 5880.7 | B |
| Bilateral total occipital lobe volume | 42666.3 ± 5434.8 | A | 45359.6 ± 5280 | A | 45858.8 ± 6133 | A | 42700.5 ± 5699.5 | A | 41479.5 ± 5934.9 | B |

*Abbreviations:* AD: Alzheimer's disease; BMI: Body mass index; CN: Cognitively normal; EMCI: Early mild cognitive impairment; LMCI: Late mild cognitive impairment; MRI: Magnetic resonance imaging; SMC: Significant memory concerns.

**Supplementary Table S8. Plasma GlycA was not correlated with CSF A/T/N biomarkers at baseline.** Spearman's rank order correlation was performed between log2-transformed A/T/N biomarkers in CSF and GlycA (log2-normalized, medication adjusted), controlling for age, BMI, APOE4, and education level, stratified by diagnosis and sex.

| A/T/N Biomarkers | Spearman rho | | | | | | | | | |
|---|---|---|---|---|---|---|---|---|---|---|
| | Male | | | | | Female | | | | |
| | CN | SMC | EMCI | LMCI | AD | CN | SMC | EMCI | LMCI | AD |
| CSF_Abeta42 | 0.151 | -0.090 | 0.042 | 0.038 | -0.002 | -0.016 | 0.038 | -0.084 | -0.074 | 0.161 |
| CSF_PTau | 0.009 | -0.159 | 0.092 | -0.093 | 0.001 | 0.041 | -0.236 | -0.047 | 0.108 | 0.151 |
| CSF_PTau/Ab42 | -0.118 | 0.007 | 0.002 | -0.092 | 0.015 | 0.037 | -0.104 | 0.069 | 0.104 | -0.028 |
| CSF_Tau | 0.075 | -0.159 | 0.124 | -0.087 | 0.003 | 0.031 | -0.175 | -0.023 | 0.079 | 0.123 |
| CSF_Tau/Ab42 | -0.088 | 0.012 | 0.021 | -0.081 | 0.001 | 0.024 | -0.080 | 0.084 | 0.090 | -0.050 |

*Abbreviations:* AD: Alzheimer's disease; A/T/N: Amyloid/Tau/Neurodegeneration; BMI: Body mass index; CN: Cognitively normal; CSF: cerebrospinal fluid; EMCI: Early mild cognitive impairment; GlycA: Glycoprotein acetyls; LMCI: Late mild cognitive impairment; SMC: Significant memory concerns.

**Supplementary Table S9. Baseline GlycA level was not associated with A/T/N biomarkers in CSF in the continuous follow-up years in participants regardless of their diagnosis at baseline.**

| Parameters | Year | p-value | | | | | Spearman rho (ρ) | | | | |
|---|---|---|---|---|---|---|---|---|---|---|---|
| | | CN | SMC | EMCI | LMCI | AD | CN | SMC | EMCI | LMCI | AD |
| CSF_Abeta42 | 1 | 0.765 | NA | NA | 0.706 | 0.209 | 0.036 | NA | NA | 0.033 | -0.164 |
| | 2 | 0.567 | 0.311 | 0.981 | 0.735 | 0.820 | -0.059 | 0.162 | 0.002 | 0.032 | 0.047 |
| | 3 | 0.367 | NA | 0.667 | 0.336 | 0.667 | -0.168 | NA | 0.500 | 0.156 | 0.500 |
| | 4 | 0.709 | 0.446 | 0.727 | 0.670 | NA | -0.050 | 0.222 | 0.054 | -0.062 | NA |
| | 5 | 0.453 | NA | NA | 0.626 | NA | -0.169 | NA | 1.000 | 0.128 | NA |
| | 6 | 0.033 | NA | 0.653 | 0.630 | NA | -0.390 | NA | -0.118 | -0.112 | NA |
| | 7 | 0.209 | NA | 0.289 | 0.600 | NA | -0.374 | NA | 0.429 | -0.400 | NA |
| | 8 | 0.600 | NA | NA | NA | NA | 0.400 | NA | -1.000 | -1.000 | NA |
| | 9 | 0.102 | NA | NA | 0.800 | NA | -0.619 | NA | NA | -0.200 | NA |
| | 10 | NA | NA | NA | NA | NA | 1.000 | NA | NA | NA | NA |
| | 11 | 0.531 | NA | NA | NA | NA | -0.262 | NA | NA | NA | NA |
| | 12 | NA | NA | NA | NA | NA | NA | NA | NA | NA | NA |
| | 13 | NA | NA | NA | NA | NA | NA | NA | NA | NA | NA |
| CSF_pTau | 1 | 0.608 | NA | NA | 0.859 | 0.207 | 0.063 | NA | NA | -0.016 | 0.167 |
| | 2 | 0.682 | 0.104 | 0.319 | 0.261 | 0.885 | 0.043 | -0.257 | 0.098 | -0.107 | -0.030 |
| | 3 | 0.535 | NA | 0.667 | 0.331 | 0.667 | 0.120 | NA | -0.500 | -0.158 | -0.500 |
| | 4 | 0.936 | 0.095 | 0.765 | 0.753 | NA | -0.011 | -0.464 | 0.047 | 0.046 | NA |
| | 5 | 0.789 | NA | NA | 0.071 | NA | 0.060 | NA | -1.000 | -0.449 | NA |
| | 6 | 0.050 | NA | 0.888 | 0.767 | NA | 0.361 | NA | 0.038 | -0.069 | NA |
| | 7 | 0.649 | NA | 0.160 | 0.400 | NA | -0.147 | NA | -0.548 | 0.600 | NA |
| | 8 | 0.800 | NA | NA | NA | NA | -0.200 | NA | 1.000 | -1.000 | NA |
| | 9 | 0.015 | NA | NA | 0.800 | NA | 0.810 | NA | NA | -0.200 | NA |
| | 10 | NA | NA | NA | NA | NA | -1.000 | NA | NA | NA | NA |
| | 11 | 0.955 | NA | NA | NA | NA | -0.024 | NA | NA | NA | NA |
| | 12 | NA | NA | NA | NA | NA | NA | NA | NA | NA | NA |
| | 13 | NA | NA | NA | NA | NA | NA | NA | NA | NA | NA |
| CSF_Tau/ab42 | 1 | 0.580 | NA | NA | 0.650 | 0.117 | 0.068 | NA | NA | -0.040 | 0.206 |
| | 2 | 0.633 | 0.062 | 0.591 | 0.809 | 0.452 | 0.050 | -0.294 | 0.053 | -0.023 | -0.154 |
| | 3 | 0.438 | NA | 0.667 | 0.620 | 0.667 | 0.150 | NA | -0.500 | -0.081 | -0.500 |
| | 4 | 0.859 | 0.267 | 0.914 | 0.553 | NA | 0.024 | -0.319 | 0.017 | 0.087 | NA |
| | 5 | 0.536 | NA | 0.200 | 0.260 | NA | 0.140 | NA | -0.800 | -0.289 | NA |
| | 6 | 0.069 | NA | 0.105 | 0.658 | NA | 0.337 | NA | 0.407 | 0.103 | NA |

| | | | | | | | | | | | |
|---|---|---|---|---|---|---|---|---|---|---|---|
| | 7 | 0.957 | NA | 0.456 | 0.400 | NA | -0.017 | NA | -0.310 | 0.600 | NA |
| | 8 | 0.800 | NA | NA | NA | NA | -0.200 | NA | 1.000 | -1.000 | NA |
| | 9 | 0.021 | NA | NA | 0.800 | NA | 0.786 | NA | NA | -0.200 | NA |
| | 10 | NA | NA | NA | NA | NA | -1.000 | NA | NA | NA | NA |
| | 11 | 0.955 | NA | NA | NA | NA | -0.024 | NA | NA | NA | NA |
| | 12 | NA | NA | NA | NA | NA | NA | NA | NA | NA | NA |
| | 13 | NA | NA | NA | NA | NA | NA | NA | NA | NA | NA |
| CSF_pTau | 1 | 0.769 | NA | NA | 0.724 | 0.677 | -0.035 | NA | NA | -0.031 | 0.055 |
| | 2 | 0.951 | 0.751 | 0.388 | 0.856 | 0.933 | -0.006 | -0.051 | 0.085 | -0.017 | 0.017 |
| | 3 | 0.658 | NA | NA | 0.865 | 0.667 | -0.083 | NA | 1.000 | -0.028 | -0.500 |
| | 4 | 0.693 | 0.091 | 0.119 | 0.926 | NA | -0.053 | -0.468 | 0.241 | -0.014 | NA |
| | 5 | 0.232 | NA | 0.400 | 0.026 | NA | -0.266 | NA | -0.600 | -0.537 | NA |
| | 6 | 0.701 | NA | 0.339 | 0.284 | NA | 0.073 | NA | -0.256 | -0.246 | NA |
| | 7 | 0.957 | NA | 0.071 | 0.600 | NA | 0.017 | NA | -0.667 | -0.400 | NA |
| | 8 | 0.800 | NA | NA | NA | NA | -0.200 | NA | 1.000 | -1.000 | NA |
| | 9 | 0.823 | NA | NA | 0.600 | NA | -0.095 | NA | NA | -0.400 | NA |
| | 10 | NA | NA | NA | NA | NA | -1.000 | NA | NA | NA | NA |
| | 11 | 0.420 | NA | NA | NA | NA | -0.333 | NA | NA | NA | NA |
| | 12 | NA | NA | NA | NA | NA | NA | NA | NA | NA | NA |
| | 13 | NA | NA | NA | NA | NA | NA | NA | NA | NA | NA |
| CSF_Tau | 1 | 0.922 | NA | NA | 0.949 | 0.534 | 0.012 | NA | NA | 0.006 | 0.082 |
| | 2 | 0.972 | 0.548 | 0.577 | 0.678 | 0.988 | 0.004 | -0.097 | 0.055 | 0.039 | -0.003 |
| | 3 | 0.855 | NA | NA | 0.888 | 0.667 | -0.034 | NA | 1.000 | 0.023 | -0.500 |
| | 4 | 0.829 | 0.274 | 0.164 | 0.602 | NA | 0.029 | -0.314 | 0.216 | -0.076 | NA |
| | 5 | 0.306 | NA | 1.000 | 0.209 | NA | -0.229 | NA | NA | -0.321 | NA |
| | 6 | 0.910 | NA | 0.338 | 0.695 | NA | 0.022 | NA | 0.248 | -0.091 | NA |
| | 7 | 0.957 | NA | 0.183 | 0.200 | NA | 0.017 | NA | -0.524 | 0.800 | NA |
| | 8 | 0.600 | NA | NA | NA | NA | 0.400 | NA | 1.000 | -1.000 | NA |
| | 9 | 0.570 | NA | NA | 0.800 | NA | -0.238 | NA | NA | -0.200 | NA |
| | 10 | 0.667 | NA | NA | NA | NA | -0.500 | NA | NA | NA | NA |
| | 11 | 0.260 | NA | NA | NA | NA | -0.452 | NA | NA | NA | NA |
| | 12 | NA | NA | NA | NA | NA | NA | NA | NA | NA | NA |
| | 13 | NA | NA | NA | NA | NA | NA | NA | NA | NA | NA |

*Abbreviations:* AD: Alzheimer's disease; CN: Cognitively normal; CSF: cerebrospinal fluid; EMCI: Early mild cognitive impairment; GlycA: Glycoprotein acetyls; LMCI: Late mild cognitive impairment; SMC: Significant memory concerns.
